# GPCR-Based Machine Olfaction On Urine Scent Surpasses PSA at Predicting Prostate Cancer

**DOI:** 10.64898/2026.07.10.26357731

**Authors:** Andreas Mershin, Claire Guest, Nikolas Stefanou, Rob Harris, Adan Rotteveel, Shannon Johnson, Kai Chau Kung, Zoi Kountouri, Howard Kivell, Elcin Zan, Clifford Gluck, Iqbal Anjum, Francesca Teasdale, Antoanela Colda, Tom Leslie, Shuguang Zhang, Claire Dowse, Keane Ong, Paul Pu Liang, Achilleas Kotsis

**Affiliations:** RealNose, www.realnose.ai, 626 Mass. Ave., 2nd Floor, Arlington, MA 02476, USA; Executive Education, MIT Sloan School of Management, 1 Main St, 9th Floor, E90, Massachusetts Institute of Technology, Cambridge, MA 02142, USA; The Osmocosm Public Benefit Foundation, www.osmocosm.org, Boston, MA, USA; Medical Detection Dogs, www.medicaldetectiondogs.org.uk, 3 Millfield Greenway Business Park, Winslow Road, Great Horwood, Milton Keynes MK17 0NP, UK; Hcyon Technology, Amsterdam, The Netherlands; Endless Frontiers Laboratory, New York University Stern School of Business, New York, NY, USA; Cleveland Clinic Nuclear Medicine, 9500 Euclid Avenue, Cleveland, OH 44106, USA; Dr. Gluck’s Wellness Center, www.cliffordgluckmd.com, 72 Sharp St Suite A-10, Hingham, MA 02043, USA; Milton Keynes University Hospital NHS Foundation Trust, Standing Way, Eaglestone, Milton Keynes MK6 5LD, UK; MIT Media Lab, 77 Mass. Ave., E14/E15, Massachusetts Institute of Technology, Cambridge, MA 02139-4307, USA

## Abstract

**Objectives:** To determine whether medical machine olfaction via tracking the activation of mammalian G-Protein Coupled odorant Receptors (GPCR) stabilized by proprietary co-polymers on a photonic MZI chip can be used to diagnose prostate cancer (PCa) via urine scent. Specifically, scent character is compared against the current diagnostic PCa screening gold-standard in the US: the serum level of prostate specific antigen (PSA). The device is an artificial "nose" sensor built on a commercial photonic platform that reads interchangeable Mach-Zehnder interferometer (MZI) chips. These chips were functionalized with a stabilised panel of mammalian olfactory G-protein-coupled receptors (GPCRs). These samples had been characterized into POSITIVE or CONTROL for PCa several years earlier by standard hospital diagnostic procedures and by trained medical detection dogs, then stored by freezing at -80 Celcius. A subset of 80 patients’ urine samples was subsequently thawed and used for training and testing the medical machine olfaction system of RealNose as an initial validation of the novel technology and methodological approach. We posed two primary research questions: (a) whether the cancer-associated odor profile would remain detectable by machine-based systems following long-term storage and with what accuracy could it be used to cluster (YES and AUC 0.79 from scent character alone), and (b) what technical and procedural requirements would be necessary to translate such a signal into a clinically useful diagnostic assay (more training samples (500 predicted to yield 0.93) and increased breadth of receptors per chip and/or more chips per device in next iteration seen as helpful).

**Design, setting, participants:** Retrospective diagnostic-accuracy feasibility study on 80 biobanked urine samples (40 PCa, 40 non-cancer; 368 sensor runs; a subset of unknown Gleason grade) from a single UK NHS urology service, the same collection used to train canine detectors.

**Main outcome measures:** Patient-level Receiver Operating Characteristic (ROC) area under the curve (AUC) under patient-grouped cross-validation with a fold-honest pooled-control reference (reconstructed from training-partition controls only); sensitivity, specificity and predictive values at pre-specified operating points; 1000-fold whole-procedure label-permutation significance; patient bootstrap 95% CIs; and leave-one-day-out / leave-one-chip-out generalisation.

**Results:** An L2-regularised linear classifier when allowed to see between three and six chips’ outcome on a patient sample extracted within-instrument AUC 0.79 (95% CI 0.69 to 0.88; 1000-permutation *p* = 0.001) from urine scent alone, exceeding this cohort’s own serum prostate-specific antigen (PSA) discrimination (AUC 0.645; itself within the population range for PSA, ≈0.67) and obtained *without a blood draw* (at the Youden point, sensitivity 0.75, specificity 0.78, PPV 0.77, NPV 0.76). This was not a plateau: AUC rose from chance at ∼30 samples, passed the serum-PSA range at ∼40, **and reached 0.79 at 80 patients (0.82 if PSA was included**, with an inverse-power fit projecting 0.93 by *n* ≈ 500 and 0.96 by *n* ≈ 1000. The discriminant was a genuine multivariate receptor pattern, independent of patient age (Spearman *ρ* ≈ 0.09; the cohort is not age-matched). So at least for these data, neither age, nor collection day, ambient humidity/temperature, or overall signal amplitude (sometimes thought of as intensity of smell) were predictive of prostate cancer status, yet the scent character was. Transfer to a new sensor chip fell to AUC 0.57 without calibration, meaning the remaining obstacles are hardware portability related rather than signal existence: much as a detection dog acclimatizes to a new setting, the system requires on-site calibration prior to use.

**Conclusions:** A genuine, confound-controlled olfactory PCa signature is recoverable from 80 samples, surpasses this cohort’s serum PSA (0.645) and exceeds the population PSA range, and improves monotonically with training-set size. We present this as a small-sample feasibility benchmark, not yet a validated diagnostic; the dominant remaining factor is training-set size, and the path to clinical-utility and improved AUC is clearly found to be a larger, multi-site, age-matched, ideally prospective training cohort. A transferable small-sample lesson is also reported: adaptive feature searches (evolutionary and self-calibrating-protocol handle search) artificially inflate cross-validation and collapse under whole-procedure permutation, whereas non-adaptive averaging survives, giving a robust scent signal obtainable from the headspace of urine samples and recorable by the RealNose device that keeps improving with expanding sample training set.

**Study at a glance:** Design: retrospective, single-batch diagnostic-accuracy feasibility study on a biobanked urine repository, reported per STARD 2015. Setting: a single UK NHS urology service (Milton Keynes University Hospital; Lancaster REC 15/NW/0527); specimens biobanked at −80◦C. Participants: 80 men undergoing urological investigation (40 biopsy-/clinically-confirmed prostate cancer, 40 noncancer controls); 368 sensor runs (mean 4.6/patient). Index test: RealNose GPCR-Mach-Zehnder interferometer urine-headspace “scent” readout *via* chips with four or eight types of mammalian human olfactory receptors functionalizing 64 available spots for Mach-Zehnder Interferometry measurements, scored by an L2-regularised linear classifier. Reference standard: histopathological (biopsy)/clinical diagnosis. Main outcome: patient-level area under the ROC curve (AUC) under patient-grouped cross-validation, with label-permutation significance and bootstrap confidence intervals.

## 1 Introduction

Serum prostate-specific antigen (PSA) remains the default first-line test for prostate cancer despite well-documented limitations: as a stand-alone discriminator for clinically-significant disease its area under the ROC curve (AUC) is only ≈0.62 to 0.68, and it drives over-diagnosis. It is also *age-dependent*, benign prostatic hyperplasia raises PSA with age, so a fixed threshold loses specificity in older and sensitivity in younger men [14, 15], and the US Preventive Services Task Current guidelines restrict routine prostate cancer screening to individuals aged 55–69 years [24, 25]. A biomarker whose discriminatory performance is *intrinsically age-independent* would constitute a superior diagnostic tool. Biological olfaction represents a candidate source of such an age-invariant signal: trained canines and laboratory analyses of urinary volatile organic compounds have achieved sensitivities and specificities in the low-to-mid 70% range on age-matched samples [1, 17, 21]. In particular, the programme led by Guest *et al.* at Medical Detection Dogs, from which the present samples were obtained, identified a urinary prostate cancer–associated signal detectable both by trained dogs and by an independent machine-learning classifier after exposure to only a few dozen specimens [1].

Building upon this work, RealNose has developed methods to stabilise olfactory G-protein–coupled receptors (GPCRs) and employ them to functionalise a commercial 64-spot Mach–Zehnder interferometer (MZI) chip [5]. This platform enables real-time interrogation of the “scent character” of urine headspace and the acquisition of quantitative recordings. The machines used in the present study were trained on the same biobanked urine samples previously used to train the medical-detection dogs that motivated this research. The primary envisioned application is as a non-invasive triage tool, requiring only a urine sample and need no blood draws, no genomic assays, and no digital rectal examination. In contrast to trained dogs, the device-based approach is amenable to large-scale deployment across multiple sites and could, in principle, be operated in any setting with access to a smartphone-based interface.

This is by necessity an extreme small-data test. Where medical-imaging artificial intelligence is trained on datasets curated at a cost of the order tens to hundreds of millions of dollars and tens to hundreds of thousands of medical images and other results while in this study the system was trained on the scent of just 80 biobanked samples. At this scale the relevant result is dual: the AUC with little training surpassed the performance of "US screening gold standard: serum-PSA" by 15 AUC points and that, importantly, a genuine, confound-free biological scent signal associated with presence of cancer is present that *improves along a learning curve*. We reason that if such improvement is observed, the route to clinical-grade performance is the conventional one of: larger, multi-site training and testing cohorts, just like any other *in vitro* diagnostic. The present study provides a confound-controlled benchmark for a medically relevant diagnostic olfactory detector trained to find prostate3 cancer in urine scent, and a transferable lesson for the small-sample, high-dimension (*p* ≫ *n*) regime; we openly report all the negative results as well presenting the methods that did not survive their controls and why they fail in the Supplementary Information section.

Summary points

#### What is already known on this topic

- Serum PSA is a weak stand-alone discriminator for clinically-significant prostate cancer (AUC ≈0.62 to 0.68) and is age-conditioned.
- Trained dogs and GC-MS analysis of urine volatiles reach low-to-mid-70s sensitivity/specificity on matched samples, establishing that a urinary-PCa scent signature exists [1].
- It has not been reported whether a small, fixed panel of human olfactory GPCRs on a photonic chip can detect this scent signature, nor how such a detector’s accuracy changes as the training set grows.

#### What this study adds

- From 80 biobanked urine samples, a fold-honest linear model extracts an age-independent, confound-audited scent AUC of 0.79 (95% CI 0.69 to 0.88; *p* = 0.001), exceeding by up to 15 AUC points the gold standard population serum PSA without a blood draw, instead from a single non-invasive scent test.
- Discrimination follows a still-rising learning curve (chance → PSA at ∼40 samples → 0.79 at 80 samples), this reframes the training-set size as the key to machine olfaction applicable to medicine.
- Cross-chip transfer falls to 0.57, localising the deployment barrier to hardware portability (addressable by on-site calibration), and a generalisable small-sample estimator lesson is documented: averaging over features does better than searching among them for the best ones.

## 2 Materials: cohort, samples, and detector

Cohort and ethics. Urine was collected under Milton Keynes University Hospital governance (Lancaster REC 15/NW/0527) from men undergoing urological investigation, including biopsy-confirmed prostate cancer across a range of Gleason grades, benign prostatic hyperplasia (BPH), and age/symptom context controls; all gave informed consent.

Sample provenance and handling. Samples were biobanked at −80*^◦^*C over 6 to 10 years at Medical Detection Dogs campus in Milton Keynes who also developed the standardized protocols used for canine training (enabling a fair machine-versus-dog comparison; freeze-thaw cycles were kept to ≤ 3). For measurement, a 1 mL aliquot was perfused with room air (60 mL/min) between one and three 60-s “sniff”/30-s “exhale” cycles with the baseline channel aspirating room air through a 0.22 *µ*m polyethersulfone membrane,with humidity and temperature recorded at each time point but not actively regulated, aside from standard human-comfort air conditioning being in operation, intermittent opening of windows, and an uncontrolled, fluctuating number of human and canine occupants in the room, thereby approximating real-world operating conditions for the envisioned practical medical olfactory device based on this technology.

Receptor panel. We built and compared two versions of the swappable MZI sensor chip, a four-receptor version and an eight-receptor version, and report the results as they came in rather than selecting one configuration after the fact. The four-receptor version carries hOR51E1, hOR51E2 (OR51E2 = PSGR), hOR17-4 and hOR2H1; the eight-receptor version carries the same four plus hOR2AG2, hOR2T7, hOR5M10 and hOR8G5. Receptors were slightly modified in their primary amino acid sequences by proprietary methods and were expressed in HEK293 cells and stabilised on the sensor surfaces using proprietary RealNose immobilisation protocols [3, 4] before being spot-printed onto the MZI arms by contractors’ nanolitter handling robot. The two versions correspond to two chip builds: the four-receptor build (batch 1) we printed each of its four receptors at two concentrations ∼6× apart as a quality control and exploratory feature, and for the eight-receptor build (batch 2) we printed each of its eight at a single concentration.

Both builds are present in the analysed cohort (two batch 1 and four batch 2 chips), so only the four receptors shared by both versions (at their high concentration) are common to every chip and cohort. Because the two builds place different receptors at the same physical spots, all features are keyed by *receptor identity* (protein, concentration), pooled over replicate spots and never over spot placement ; the full chip protein population, layout collisions and concentration design are given in SI-G.

As many of the cohort were read on all available chips to treat that heterogeneity as an asset rather than a nuisance. Each specimen was exposed to as many distinct receptors and as many independent devices as logistics allowed, and a patient’s run-level scores were then averaged, so that per-spot and per-device noise canceled. A four-receptor chip contributes those four receptors; an eight-receptor chip contributes the same four plus four more. A receptor that a given chip does not carry is imputed to the pooled-control baseline, so it adds no discriminative signal and a chip is never penalised for lacking it. Because features are matched by protein identity, mixing the two builds never averages one receptor against another; it only widens the receptor variety seen per patient, which is what helps when there is only a small cohort to average out noise without over-fitting.

Label and clinical characteristics. The label is the clinical POSITIVE/CONTROL diagnosis, which is potentially imperfect in the ways we report here. (Table 1). The 40 controls are a symptomatic urological work-up population (22 non-malignant disease, 6 no evidence of malignancy, 12 unspecified; organ prostate 9, bladder 10, kidney 1, testes 1), not healthy volunteers, and the two arms are *not* age-matched (cancer 67.7±8.4 y vs control 51.8±18.7 y, the cohort-composition confound of §6). Cancer status is taken from the clinical/biopsy diagnosis independently of Gleason grade: of the 40 positives, 37 carry a parseable Gleason score (6/7/8/9 = 16/16/3/2) and 3 do not, and those three are retained as POSITIVE on the strength of the clinical diagnosis rather than dropped. Both the comorbid control arm and the imperfect label depress any cancer-versus-non-cancer AUC.

**Table 1:** Clinical and demographic characteristics of the analysed cohort (80 patients, 368 runs). Percentages are of the column arm.

| Characteristic | Cancer ($n = 40$) | Control ($n = 40$) |
| --- | --- | --- |
| Age, mean ± SD (years) | $67.7 \pm 8.4$ | $51.8 \pm 18.7$ |
| Age range (years) | 39 to 82 | 19 to 88 |
| Sensor runs, mean per patient | 4.6 (range 3 to 6; 368 total) |  |
| *Reference-standard basis* |  |  |
| Biopsy/clinically-confirmed cancer | 40 (100%) |  |
| Non-malignant urological disease |  | 22 (55%) |
| No evidence of malignancy |  | 6 (15%) |
| Unspecified |  | 12 (30%) |
| *Gleason grade (cancer arm)* |  |  |
| Gleason 6 | 16 |  |
| Gleason 7 | 16 |  |
| Gleason 8 | 3 |  |
| Gleason 9 | 2 |  |
| No parseable Gleason | 3 |  |
| *Organ under investigation (control arm)* |  |  |
| Prostate / Bladder / Kidney / Testes |  | 9 / 10 / 1 / 1 |
| BPH explicitly noted |  | 2 |

Analysed cohort. These specimens derive from a larger institutional recruitment programme (1110 patients assessed, 901 consented; plus 260 self-referred healthy volunteers), whose recruitment flow and differential patient-versus-volunteer recruitment are documented in SI (Fig. SI-1); the controls analysed here are predominantly the symptomatic work-up patients, not those healthy volunteers. To hold the measurement instrument fixed, cross-batch and cross-chip variation being precisely the confound this paper isolates (§6, §8), all analyses use a single verified sensor dataset (368 runs). Labelled by clinical diagnosis (POSITIVE/CONTROL), this dataset contains 80 patients (40 cancer, 40 control), on which all reported results are computed. Runs vary in recorded length, but the discriminative signal lies in the early analyte phase of each trace (§3); specimens whose acquisition stops before the late phase therefore carry the same information as the rest and are retained, so no patient is excluded on acquisition length. The full participant flow is given in the SI (Fig. SI-2).

## 3 Methods

Feature construction. Each of 64 spots left an analyte-phase trace, expressed as a deviation from its own baseline and is summarised by six statistics (maximum, minimum, mean, final value, range, and slope). These six values across all populated spots (∼348 spots × 6) gives the 2088-dimensional feature vector per run, to which the pooled-control referencing of the next paragraph is then applied. Pseudocode and workflow flowcharts for the full pipeline are given in SI-Code.

Fold-honest virtual pooling. Rather than use each sensor’s raw output, we express every reading as a *deviation from normal urine*: we compute a reference vector, the mean response of the non-cancer (control) samples, and subtract it from each patient’s measurement, so the features describe how a sample differs from a locally-defined control. This “virtual pooled control” is the digital equivalent of physically pooling a batch of local negative-control urine samples into one reference measurement. Because it is rebuilt from whichever controls are on hand on the day and in the room tests are about to take place this method self-calibrates against device-to-device and site-to-site drift (RealNose, Inc. virtual-poolingmethod, 4). To keep the evaluation leak-free, the reference and the feature scaler are estimated on the training-partition controls only and then applied to held-out patients, with a leave-one-out correction that prevents any test sample from entering its own reference.

Estimators. (a) A reciprocally-conditioned weak-learner “swarm” with a Machine Performance Index (the biomimetic origin; SI-B); (b) an L2-regularised logistic classifier (primary): a gently-restrained linear vote across all receptors, the "ridge penalty" forces modest influence to be spread across many features rather than concentrated on a few, which is correct when the signal is diffuse (§5). It performs no feature search and has a single hyper-parameter, so there is nothing to over-search; this is why it survives permutation where adaptive methods do not (§6).

Significance and stress tests. Patient-grouped leave-one-patient-out (LOPO) and 5-fold cross-validation; 1000-fold label permutation of the *entire* pipeline; patient-level bootstrap confidence intervals; and leave-one-*day*-out (LODO) as a detector-drift stress test.

Debiasing. Baseline-drift removal, per-day quantile normalisation, covariance alignment (CORAL), and a domain-adversarial variant were evaluated as cross-session corrections (§8, SI-A; cf. prior debiasing work 2); none completely recovered cross-instrument transfer.

## 4 Results I: discrimination and its dependence on training-set size

The central result is dual: a) the accuracy at training sample size 80 is already surpassing PSA AUC by 15 points and b) this accuracy grows steadily as the training set grows. Sub-sampling the 80-patient cohort under identical patient-grouped cross-validation, scent-only discrimination rises with cohort size (Fig. 2): from chance at ∼16 to 24 samples, through the population serum-PSA level (≈0.67) at ∼40, to AUC 0.79 (95% CI 0.69 to 0.88; 1000-permutation *p* = 0.001) at 80, from urine scent alone and 0.82 if PSA value was also allowed to be included. This value is a patient-level figure obtained by averaging each patient’s repeat measurements (mean seen by 4.6 chips each); a single reading gives only 0.69, and it takes about four to five repeats to reach 0.79 (§8). The curve does not saturate; an inverse-power fit AUC(*n*) = 1 − 3.3 *n^−^*^0.63^ projects ≈0.86 at *n* = 150 and over 0.93 *n* ≈ 500 and at 0.96 by *n* ≈ 1000 (extrapolation beyond the observed range). The dominant remaining factor is found to be training-set size (within-instrument; cross-site deployment can be done cold but improves with the on-site calibration of §8).

**Figure 1:**
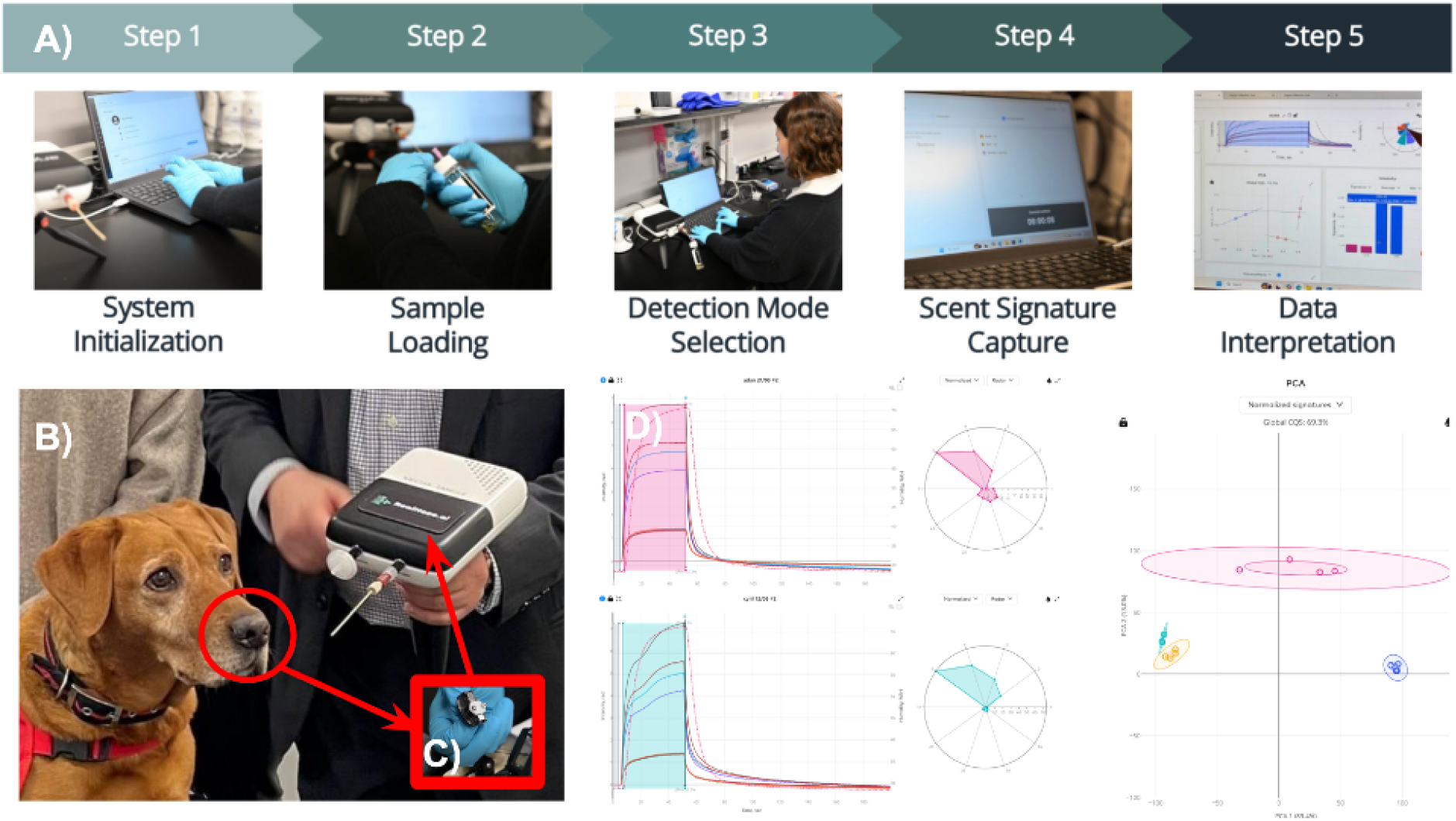
RealNose sensor and training/testing workflow on urine scent for prostate cancer diagnosis inspired by medical detection dogs. A) The five steps of training and detection via RealNose sensor. B) To-scale comparison of Florin the prostate cancer biodetection dog, the RealNose olfactory receptor based sensor platform and the C) Two-path, commercially available quick-exchange biophotonic MZI with 64 spots populated by bioprogrammable RealNose olfactory receptors. D) Exemplary data obtainable from typical scent collection runs on two volunteers’ urine.

**Figure 2:**
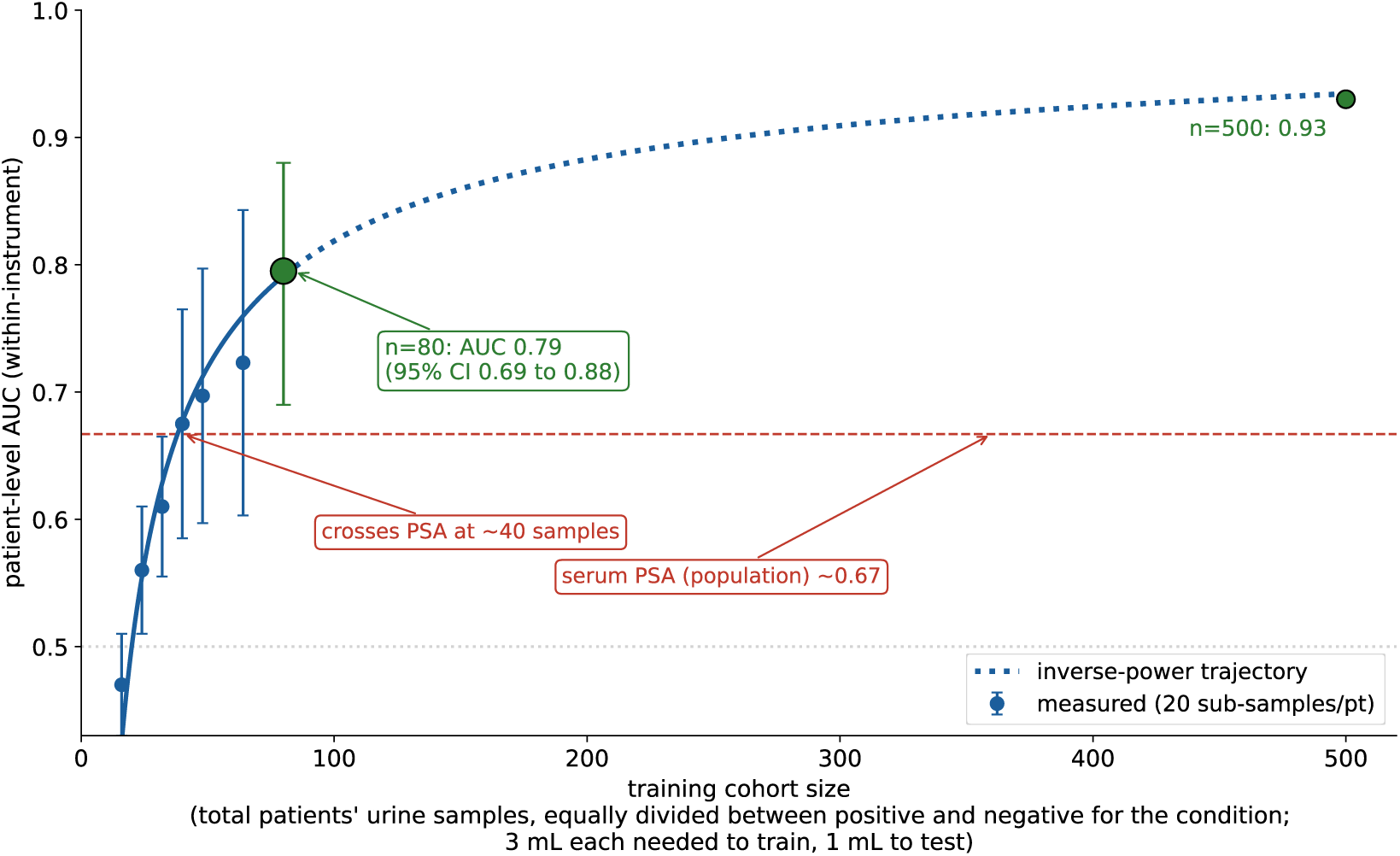
Patient-level discrimination versus training-cohort size. Patient-level AUC versus training-cohort size (analysed dataset, 20 random samples per point). AUC rises from chance (∼30 samples) past the population serum-PSA line (≈0.67, at ∼40 samples) to 0.79 when using 80 samples, with an inverse-power extrapolation to past 0.93 at n=500 and 0.96 by *n* ≈ 1000 predicted. Within-instrument / same-sensor; cross-site transfer benefits from on-site calibration (acclimatization step involving sniffing a known negative baseline sample) (§8).

The reported classifier is a plain L2-regularised logistic model with no feature search; a biomimetic reciprocal “swarm” (a machine analogue of the *Drosophila*Performance Index [18–20]) motivated the work but does not beat it (0.73 vs 0.79; SI-B). Interestingly, fusing scent with serum PSA gives a small but genuine gain (0.79 → 0.82; PSA-alone is at 0.645 for this set), so scent is not redundant with PSA i.e. we are not sniffing out (a proxy for) PSA. Adding age raises the apparent AUC further (scent+PSA+age makes AUC=0.89) but since that depends on the cohort’s age imbalance (§6) it is not claimed as part of the record AUC; we report scent-only (0.79) as the primary, age-independent result and scent+PSA =0.82 as an interesting fusion with a widely available blood diagnostic priced at around 20USD which by itself only reaches AUC of 0.67. Noting also that applying a dog-like conformal-abstention rule (deciding on only the most confident samples) reaches AUC of ∼0.94 at 50% coverage. Because an AUC alone does not convey clinical behaviour, Table 2 gives two pre-specified operating points: balanced (Youden) operation yields sensitivity/specificity in the 0.70-0.80 range, and forcing sensitivity to 0.90 costs specificity (0.40) -as is the expectation.

**Table 2:** Scent-only operating points (patient-level, patient-grouped cross-validation; predictive values at the cohort prevalence of 50%). AUC 0.79.

| Threshold | Sensitivity | Specificity | PPV | NPV |
| --- | --- | --- | --- | --- |
| Youden-optimal | 0.75 | 0.78 | 0.77 | 0.76 |
| High-sensitivity ($\geq 0.90$) | 0.90 | 0.40 | 0.60 | 0.80 |

## 5 Results II: the nature of the discriminant

The discriminant is a genuine *multivariate relative-magnitude pattern* across olfactory receptors, not overall response amplitude: a total-magnitude scalar performs no better than chance (AUC 0.54). We recognize our signal as being diffuse rather than sparse (poetically appropriate to a lingering smell permeating many samples concept) because nested feature selection degrades performance monotonically (top-10 → 0.50; top-100 → 0.66; all-2088 → 0.79), so no low-dimensional “handle” suffices, which explains why adaptive handle searches fail (§6). Consistently, no single receptor discriminates all on its own (all ≤0.53), and discrimination improves as more receptors are combined, reaching its best with the full panel so it is breadth across receptors, not any privileged receptor that allows this discrimination to happen (SI-F) and it stands to reason adding more receptors to each chip will keep improving the discrimination.

We also see that the response is genuine receptor binding, not a bulk or instrument artifact: a six-fold increase in printed receptor produces a near-constant two-fold rise in signal across all four receptors (2.00 ± 0.14; the saturable Langmuir/Hill signature [22] that a bulk effect cannot produce), (SI-H). Because this high/low ratio is so tightly conserved, it also serves as a built-in, calibrant-free quality check: a spot pair that drifts outside the narrow band flags a potentially denatured, defective or fouled receptor. The concentration *ratio* itself carries no cancer information (AUC 0.50), so the discriminant lies in the cross-receptor pattern; the full mechanism and its controls are given in SI-H.

## 6 Results III: confound and nuisance controls

Age (critical). The cohort is not age-matched (cases 67.7 y, controls 51.8 y). Age alone predicts the label at AUC 0.76, but this is cohort composition, not biology: the scent signal itself does not encode age (Spearman *ρ* ≈ 0.09; MAE 15 y = population-mean guessing). Spearman’s *ρ* is a rank correlation ranging from −1 to +1, where 0 means no consistent relationship and ±1 a perfect one; at *ρ* ≈ 0.09 a model given only the scent cannot recover a patient’s age (its error, 15 years, is no better than guessing the group mean). We therefore *exclude* age; the reported 0.79 is age-independent. Combining scent+PSA+age reaches 0.89, but its excess over 0.82 uses the age artificial confound and can not be claimed as a real improvement.

Disease burden (does "more cancer" smell "more like" cancer?). A reasonable expectation is that higher-grade or higher-volume malignancies may be more readily detectable if higher-grade cancer is associated with a greater perceived odor intensity. This presupposes that increased olfactory intensity corresponds to a larger quantity and/or stronger binding affinity of volatile organic compounds emitted by higher-grade disease. Moreover, it can be hypothesized that comorbid pathological conditions could augment a more generic “cancer” odor signature. Neither holds in these data (SI-F) although with larger datasets such trends may be indeed found. Here however we see that the per-patient score is uncorrelated with Gleason sum (Spearman *ρ* = −0.07), positive-core fraction (*ρ* = 0.05) or serum PSA (*ρ* = −0.15; all *p >* 0.37); among controls, those with documented non-malignant disease were, if anything, scored *lower* than disease-free controls (no control in this cohort carries a cancer, so the strong “other-cancer” hypothesis is untestable here). Detection via RealNose sensors is therefore, at this level of training, burden-*independent*: the readout registers the presence of a prostate-cancer scent character rather than grading how much disease is present. This is a hypothesis-generating, single-batch observation (*n* ≈ 37 to 40 with burden fields), but its direction matters: it reinforces that the dominant factor in performance is training-set size (§3), not disease severity, and it cautions against expecting easier detection in advanced disease and spurs optimism that this method might, like the trained canines offer early detection.

Collection day and sensor chip (the acclimatization axis). The three measurement days are label-balanced (41/51/50% positive), so day is not a label proxy, day predicts the cancer label at only AUC 0.53 (95% CI 0.42 to 0.65; Fig. 3), even though day is predictable from the raw signal (AUC 0.88) so the drift is real but disease-uncorrelated. Within-instrument discrimination is 0.79; leave-one-day-out is 0.57; leave-one-*chip*-out is 0.56. This descending gradient, the more the evaluation is forced to be independent of the measurement apparatus, the closer to chance, is the central translational finding: the biological signal exists and is highly significant, but a model trained on one chip/session does not transfer unchanged to new hardware. This is the machine analogue of a detection dog (or clinician) needing to acclimatize to a new facility, and it motivates on-site control-calibration (§8) rather than a fixed factory model.

**Figure 3:**
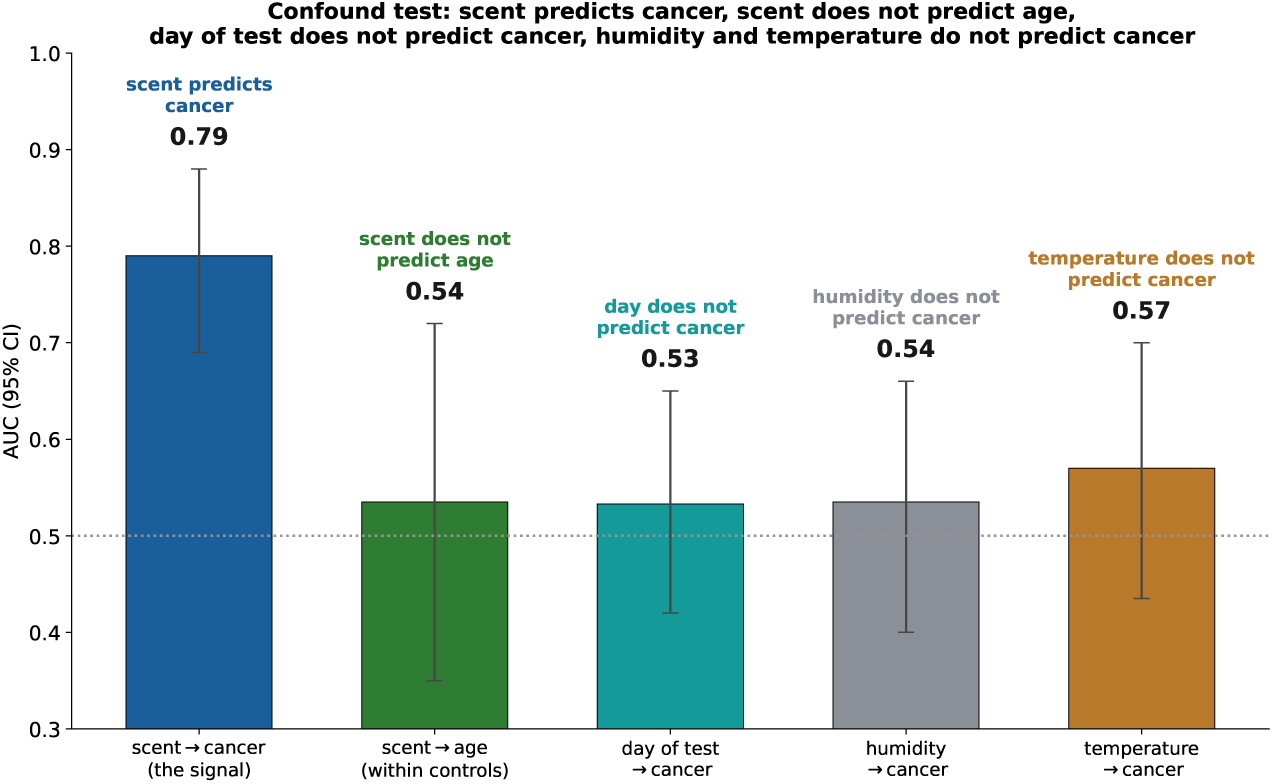
Confound test: scent predicts cancer, scent does not predict age, day of test does not predict cancer, humidity and temperature do not predict cancer. Patient-level patient-level AUCs with 95% bootstrap confidence intervals (error bars). The scent readout discriminates cancer from control (0.79) but does not predict patient age, evaluated *within controls* to avoid the cohort’s age-cancer correlation, scent reads old-vs-young at 0.54 (95% CI 0.35 to 0.72; consistent with the age-regression *ρ* ≈ 0.09). The day of test (0.53, CI 0.42 to 0.65; the three days are label-balanced, 41/51/50% positive), ambient humidity (0.54, CI 0.40 to 0.66) and temperature (0.57, CI 0.44 to 0.70) likewise do not predict cancer. Patient age is not shown here because it *does* predict the label (AUC 0.77) purely through cohort composition (cases 67.7 y vs controls 51.8 y; *not* age-matched); age is a confound and is therefore excluded from the model, so the reported discrimination is age-independent (§6). (Cross-day *generalisation*, a separate matter, is addressed by the leave-one-day-out analysis of §6, §8.)

Humidity/temperature. Neither ambient humidity (patient-level AUC 0.54, 95% CI 0.40 to 0.66) nor temperature (0.57, CI 0.44 to 0.70; Fig. 3) predicts the cancer label, so neither is a label confound. Nor is the discriminant explained by humidity: both subtractive (regress-out) and multiplicative (gain/interaction, 126 features) recodings of the signal around humidity destroy rather than concentrate the signal (recoded AUC ≈0.38), i.e. the discriminant is neither humidity-orthogonal nor humidity-coupled. Humidity is a genuine nuisance modulator, but recoding the signal around it only discards information, a tested-and-rejected confound hypothesis.

## 7 A methodological caution: searching for the best features artificially inflates accuracy

Two of the methods we evaluated search for the most discriminating combination of sensor features: one by a guided search (the self-calibrating protocol, SCP [6–8]), the other by an evolutionary algorithm. Both reached an apparent AUC of 0.82 to 0.83, but this gain was an artefact. When we re-ran the *entire* procedure many times on randomly permuted labels, the search still returned an AUC of 0.84 (*p* = 0.63), performing as well on permuted labels as on the real ones, meaning that the apparent gain reflects the ability of the search to overfit rather than the results being associated with a real biological scent signal for cancer. We safeguarded against this applying the significance test to the whole search, not only the final model, and this led us to gravitate towards the reported methods that tend to average over all features rather than selecting a small, high performing subset. In other words, when available features greatly outnumber patients, selecting among them tends to overfit, whereas averaging across them is more robust. A single-receptor ratio illustrates the same effect: it appeared strong on the data from which it was selected (0.86) but fell to chance (0.36 to 0.50) under this evaluation. Table 3 lists every method evaluated under this protocol; averaging across all receptors (the L2-logistic model, 0.79) performs best, and every attempt to compress the signal into a few features performs worse.

**Table 3:** Method comparison on the study cohort. All AUCs are patient-level under patient-grouped cross-validation; “honest” denotes the value that survives whole-procedure label permutation. Adaptive searches are shown with their in-sample number and their permuted null.

| Estimator / coding | Honest patient AUC | Note |
| --- | --- | --- |
| L2-logistic (all receptors) | 0.79 | primary; no feature search |
| Reciprocal swarm / MPI | 0.73 | biomimetic origin (SI-B); does not beat linear |
| FlyHash sparse expansion | 0.66 | real but < logistic |
| Total-magnitude scalar | 0.54 | signal is not overall amplitude |
| Concentration ratio | 0.50 | dose-response carries no cancer info |
| Primacy / onset-rank coding | $\approx 0.50$ | chance |
| Nested top-10 / top-100 features | 0.50 / 0.66 | signal is diffuse, not sparse |
| Single-receptor ratio (hOR51E1/hOR17-4) | 0.36 to 0.50 | in-sample 0.86; does not reproduce under permutation |
| Humidity residual / gain (126 feats) | $\approx 0.38$ | tested-and-rejected confound |
| Age alone | 0.76 | cohort-composition confound; *excluded* |
| Serum PSA alone | 0.645 | this cohort |
| *Adaptive (inflated) searches* |  |  |
| SCP random-handle search | $0.83 \rightarrow 0.835$ ($p=0.63$) | collapses under permutation |
| Evolutionary handle optimisation | $0.82 \rightarrow 0.84$ ($p=0.63$) | similar procedure, same collapse |

Other estimators evaluated and not exceeding L2-logistic (L1-logistic, LDA, PLS-DA, linear/RBF SVM, random forest, extra-trees, gradient boosting, *k*-NN, naive Bayes, stacking, SPRT sequential sniffing, CORAL/quantile drift-removal, MFP density, noise augmentation) are listed in SI-C.

## 8 Cross-session generalisation and on-site calibration

Leave-one-day-out AUC is ≈0.57 and leave-one-*chip*-out is ≈0.56; the cancer discriminant direction rotates across sessions (cross-day cosine −0.15), and covariance alignment (CORAL), day-predictive-feature removal, and per-day quantile normalisation do not recover it, with only three measurement days a drift model is under-determined. The biological signal exists and is highly significant, but a model trained on one chip/session does not transfer unchanged to new hardware. This is the machine analogue of a detection dog (or clinician) needing to acclimatize to a new facility.

On-site calibration by virtual pooling. The concrete acclimatization step is a*site-specific pooled-control reference*: at each deployment site a small panel of locally-run non-cancer (control) urines is measured on the local chip, their deviation-bearing sensograms are averaged into a local reference, and every subsequent unknown is expressed as a deviation from that local reference before scoring. This is the analysis-side analogue of physically pooling local negative-control urine into a single reference measurement, and it uses only the labels of the local *controls*, never the unknown’s label, so it is leakage-free. We contrast two regimes on the study cohort (Fig. 4): *cold* transfer (a fixed model applied absent local calibration / acclimatization, referenced only to training-site controls) and *calibrated* transfer (each session referenced to its own local controls).

**Figure 4:**
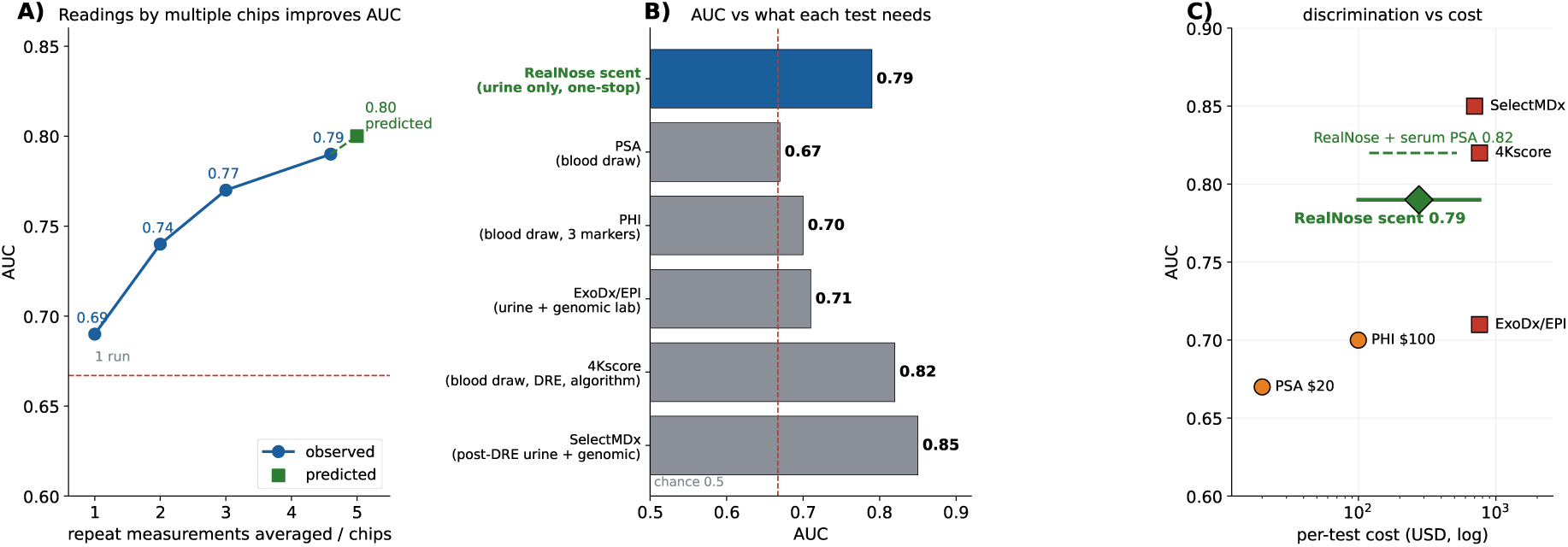
All panels plot AUC, the area under the receiver-operating-characteristic curve for clinically-significant prostate cancer (Gleason 3+3 or above) at the patient level: the probability that a randomly chosen cancer patient is scored higher than a randomly chosen control, where 0.5 is chance and 1.0 is perfect discrimination. (A) Reading by more than one chip: a single chip gives 0.69; a second and third chip’s reading raise it to 0.74 and 0.77, and each patient’s full set (mean 4.6 runs, across ∼3 to 4 chips) reaches 0.79, with 0.80 projected at five runs, adding a known PSA bumps up final AUC to 0.82. (B) AUC versus what each test requires: RealNose (urine scent 0.79 + PSA = 0.82) exceeds population PSA (0.67), PHI (0.70) and ExoDx/EPI (0.71) only the $600 to 760 blood/genomic panels (4Kscore 0.82, SelectMDx 0.85) score higher; the label beside each bar states what that additional tests each method needs to reach stated AUC (RealNose requires one urine sample, no blood draw, no digital rectal examination, no genomic send-out, if PSA available it adds +3AUC points making AUC 0.82). (C) Discrimination versus per-test cost (Medicare CLFS benchmark, USD, log axis): from a single non-invasive urine test the scent readout (0.79) attains discrimination comparable to validated adjunctive assays across the $100 to $760 CLFS range, and adding serum PSA gives a modest further gain (0.82). Comparator AUCs are literature values from larger validated cohorts; the fair claim is comparable discrimination at a fraction of the data, cost and invasiveness. No price is assigned to the urine-scent test by RealNose yet and it is not available to the public.

The two regimes, together with multi-measurement voting and the observed (up to four) and extrapolation to five measurements, are summarised in Fig. 4.

On this cohort, virtual-pooling calibration moves cross-session AUC only marginally (cross-chip 0.51 → 0.53; cross-day 0.57 → 0.58): with only 80 samples and few chips, the local-control reference cannot yet model the drifts, so we do *not* present on-site calibration as the near-term priority only as a potential improvement once the base AUC and other issues standing between medical olfaction and modern clinical diagnostics are cleared. Of the interventions we tested, the two that demonstrably move AUC performance upwards are both forms of *more information*: more training patients, which raises the within-instrument ceiling along the learning curve of Fig. 2, and more repeat measurements of the same patient by different chips, and more receptors per chip all of which averages out single-shot noise. Local acclimatization, by contrast, buys a few points at most on present evidence. This ordering has a precise biological parallel. A medical-detection dog also acclimatizes to a new room, handler and ambient conditions before working a fresh site, a AUC-increasing step, yet no amount of acclimatization substitutes for the hundreds of rewarded training exposures that make the animal competent in the first place. For the machine as for the dog, and indeed as for a large language model, or human physician the dominant determinant of performance is the quantity and quality of training experience; site adaptation is a second-order correction layered on top.

### 8.1 Repeatability: a single chip’s reading is improved by a second and third chip

Every patient was measured multiple times (3 to 6 runs, mean 4.6). A single chip’s reading is sharpened by adding a second and third chip’s opinion. Individual runs are noisy, a random pair of a patient’s own runs disagrees on the binary call 40% of the time (within-patient score standard deviation 0.23), and pooled run-level AUC (scoring every run separately) is ≈0.64, but this averages out as chips are combined: patient-level AUC rises 0.69 (one chip), 0.74 (two), 0.77 (three), to 0.79 at the full ∼4.6 (Fig. 4). Combining several chips’ readings is therefore itself a practical form of calibration, and because multiple chips can be built into one instrument this is a hardware feature (does not mean repeat visits or repeat experiments, just one urine seen by several chips/receptor panels). (This is a within-instrument gain; it does not by itself resolve the cross-chip transfer of the preceding paragraph, which additionally requires local-control acclimatization to perform fully.)

## 9 Comparison and value

At AUC 0.79 from urine alone, the detector exceeds this cohort’s own serum PSA (computed to be 0.645) and the population PSA range (≈0.67), and sits within the band of validated multi-analyte assays (PHI, ExoDx/EPI, 4Kscore, SelectMDx), while requiring no blood draw, no genomic assay and no digital rectal examination the others depend on (Fig. 4). Those comparator AUCs come from large externally-validated cohorts, whereas ours is small-*n* cross-validation; the defensible claim is therefore *comparable discrimination at a fraction of the data, cost and invasiveness, pending prospective validation*. Benchmarking cost by the reproducible U.S. Medicare Clinical Laboratory Fee Schedule (CLFS) [13], the three adjunctive panels, ExoDx/EPI (0005U; 10), SelectMDx (0339U; 11) and 4Kscore (81539; 12), each reimburse at $760, versus ∼$20 for serum PSA and ∼$100 for the Prostate Health Index; At present, no pricing has been established for the RealNose urine scent test, and the assay is not yet commercially available, as it has not yet been tested in multi-site clinical validation trials. The intended value proposition is a triage tool based on a single urine specimen that achieves diagnostic discrimination comparable or exceeding performance of existing methods, but bearing all the advantages of being entirely non-invasive (urine scent) and yet riding on available clinical laboratory modality (urine samples) potentially unlocking a substantially lower cost for mass screening. The technology is not yet positioned to substantiate claims of superior raw area-under-the-curve (AUC) performance relative to all alternative approaches. Nonetheless, canine olfactory detection has repeatedly demonstrated superior sensitivity and specificity compared to currently available clinical tests for certain conditions, and its high diagnostic accuracy and rapid response times continue to serve as the primary conceptual and performance benchmarks guiding this development.

Adding serum PSA. Where a blood sample is available, combining the scent score with serum PSA raises patient-level AUC from 0.79 to 0.82 (Fig. 4C, dashed line). PSA contributes partly-independent information rather than duplicating the scent signal, PSA alone reaches only 0.645 in this cohort, so an inexpensive and already-routine blood test adds a small but genuine increment at no extra procedural burden. This places the combined test (0.82) level with the 4Kscore and just below SelectMDx (0.85) without needing a genomic sequencing step. We report scent-only (0.79) as the primary result and scent+PSA (0.82) as the realistically practical combined figure.

Fit with UK NHS priorities. These trade-offs map onto stated NHS priorities. Prostate cancer is the most common cancer in men in the UK, with about 57,900 new cases and 12,300 deaths each year [27] and the second most common cancer in men worldwide [16], yet only around half are diagnosed at an early, more-curable stage (53% at stage I or II in England in 2022 27), against the NHS Long Term Plan ambition to diagnose 75% of cancers at stage 1 or 2 by 2028 [26]. Delivering high-quality diagnosis while controlling cost calls for triage that catches more cancers early, reducing under-diagnosis, without inflating the over-diagnosis and over-treatment that follow from PSA’s limited specificity, and without the biopsy-related morbidity that a PSA-driven referral can carry. A low-cost, entirely non-invasive urine test that matches or exceeds this cohort’s serum PSA (0.645; population range ≈0.67), and adds partly-independent information when combined with it, is a natural candidate first-line filter: it could help prioritise who proceeds to multiparametric MRI and biopsy while sparing others an invasive work-up, aligning earlier, higher-quality diagnosis with the value and cost-control agenda. We present this as a direction the present feasibility data motivate.

## 10 Discussion

### 10.1 Statement of principal findings

From 80 biobanked urine samples, some of unknown Gleason grade and labeled only as cancer or control, a regularised linear model gives a permutation- and bootstrap-validated AUC of 0.79 from urine scent alone (95% CI 0.69 to 0.88; *p* = 0.001), first matching (at 40 training samples) and then exceeding (AUC 0.79 at 80 traning samples) the population discrimination of serum PSA (0.645 for this cohort) from a single non-invasive test (no blood draw, no genomic assay, no digital rectal examination required). We highlight three further observations as noteworthy emergent insights from this result: a) the signal corresponds to an age-independent multivariate receptor pattern (§4–§6); b) discrimination performance continues to improve as the training-set size increases and has not yet saturated at 80 samples (§3); and c) in our progress toward a practical diagnostic, the primary bottleneck now appears to be cross-instrument transfer rather than on-site raw performance, which can be enhanced through more chips per instrument and or more receptor per chip and device acclimatization / calibration. Our observations and conclusions are supported by quantitative metrics and by demonstrated repeatability across multiple MZI reading base stations without re-training, with the role of acclimatization and reproducibility explicitly examined in (§8).

### 10.2 Strengths and weaknesses of this study

The main strengths are a deliberately conservative analysis and full disclosure. Throughout our work every model is trained and tested on separate patients (patient-grouped cross-validation), and the control reference and feature scaling are computed only on the training patients, so no information leaks from train to test. Significance is assessed by permuting the labels through the whole procedure, and uncertainty by patient bootstrap confidence intervals. Generalisation is tested by holding out a whole measurement day or a whole sensor chip at a time. Finally, every confound we could identify was tested and, where it explained the label, excluded (§6). The result is a *performance floor* : we cannot verify that every specimen carried the target odour, since some tumours may be volatilome-silent, storage may rework the profile, and a biopsy-positive the dog cannot smell is, for a scent learner, a mislabelled example that only depresses the ceiling. One reassurance follows from the shared provenance: because these specimens come from the dog-characterised biobank, we have qualitative, programme-level assurance that the positives at least at the time of testing which was potentially up to 8 years ago still carried a genuine, canine-detectable odour and were not consistently scored blank by the dogs, an independent check that a real odour was present, though the dataset holds no per-sample dog verdicts to regress against. The weaknesses bound the claim: a single retrospective batch of *n* ≈ 80; a symptomatic, non-age-matched control arm (Table 1); a coarse binary label; three measurement days; and unproven cross-site transfer. We make these explicit in a QUADAS-2 self-assessment (SI-E2), rated *high* risk of bias for patient selection and *unclear* for index-test blinding, declared, not left to be inferred, and each the target of a prospective study currently under design.

### 10.3 Comparison with other studies

The result is consistent with prior work. Trained dogs and laboratory analysis of urine volatiles reach AUCs in the 0.70-0.80 sensitivity/specificity on matched samples [1, 17, 21]; the Guest *et al.* programme, from which these very samples derive, recovered a urinary-PCa signal, by both dog and an independent machine classifier, after training on only 37 specimens [1]. That an independent sensing modality (mammalian GPCRs on a photonic interferometer, a different transducer and a different learning method) lands in the same 0.70-0.80 range at the same small scale is convergence of *conclusion*, not of technical implementation, and strengthens the inference that a genuine, learnable urinary-Prostate cancer specific scent character signature exists. Against clinical comparators, scent (0.79) exceeds this cohort’s own serum PSA (0.645) and the population PSA range (≈0.67), and sits within the band of validated multi-analyte adjunctive assays (PHI 0.70, ExoDx/EPI 0.71, 4Kscore 0.82, SelectMDx 0.85), which are validated in far larger cohorts but require blood, a genomic send-out and/or clinical inputs and cost over 600USD each without counting the additional test and logistics costs of multi-test protocols carry (§9).

### 10.4 Meaning of the study: possible mechanisms and clinical implications

AUC as shown in Fig. 2 rises with training-set size: the machine performs no better than chance when trained on below ∼40 patients, it is crossing PSA performance at above 40, and is still rising at 80 training patient samples. The practical implication is that the main factor on performance is the amount of training data, so clinical-grade accuracy should follow from a larger, multi-site, age-matched cohort rather than from new physics or a new algorithm. This is the same data-centric behaviour seen across contemporary machine learning, where accuracy tends to follow empirical scaling laws in the size and diversity of the training set and where models improve as they see more examples; a molecular sensor read by a linear classifier is subject to the same forces, which is why we treat training-set size, rather than the sensor or the algorithm, as the binding constraint. The readout is consistent with genuine receptor binding (saturable and receptor-dose-dependent, *k* = 0.39; §5), and its discriminant is distributed and low-amplitude, spread across different olfactory receptors, insensitive to overall response magnitude (if trying to discriminate on total-magnitude we only managed an AUC of 0.54), and independent of age and disease burden. A signal carried by many varying volatiles all detected by receptors that are differently tuned to each in a complex plume rather than a few dominant markers would be expected to remain detectable under heavy aqueous dilution and across markedly different diets, regimes in which canine detection of this odour is reported to persist; we present this as a qualitative consistency only. A second, complementary route to higher accuracy operates alongside training-set size: individual runs are noisy (test-retest disagreement ≈40%), but this noise averages out, so combining the readings from about four chips per patient reaches the observed 0.79 for using 4.6 chips on average per patient and the projected 0.80 or above if we built a machine with 5 chips instead of one (Fig. 4A). Because multiple chips can be integrated into a single instrument, this averaging is a property of the hardware rather than a requirement for repeat visits, and supports an anticipated operating AUC above 0.80 with the present design. Two implications follow. Even at this small training scale, the test already exceeds this cohort’s serum PSA (0.645), and the population PSA range, from a single non-invasive urine sample, so a deployable triage is almost attainable now: rapid, age-independent, and requiring no blood draw, genomic assay or digital rectal examination. More consequentially, the learning curve has not plateaued, and the more valuable path is to raise discrimination further with larger and more diverse training cohorts (targeting 1000 samples balanced 50:50 between PCa+ and PCa-), improve the number of receptors per chip, increase the number of chips per test to 5, deploy at scale, and allow the model to keep improving as data accumulate. This will unlock the scaling and transfer-learning available to machines but not to trained animals.

## 11 Conclusion

An AUC of 0.79 from urine scent alone (0.82 if PSA knowledge is also allowed), is age-independent, confound-audited, exceeding the population serum PSA gold standard, and reaching 0.94 on the confident half of patients. These results were obtained from a small, dog-shared cohort whose learning curve continues to rise. We present it as a small-sample feasibility benchmark rather than a validated diagnostic, with a transferable methodological point (in high-dimension, small-sample data, averaging over features is more reliable than searching among them) and a locked prospective design (§**??**) to confirm it.

While AUC of 0.79 is 12-15 AUC points better than the current US prostate cancer screening "gold standard" of serum PSA we still see this as a modest AUC for any screening tool. Especially given the extraordinarily small dataset used (40 negative and 40 positive patients’ samples only) we find this result consequential rather than incremental: while AUC is certain to rise with more training samples, this results already brings dog-like scent performance, and dog-like training, within reach of machines, while adding what machines uniquely provide: the beginnings of transfer learning, scaling across many sites, and continual improvement from accumulated data. Because the sensing elements are the same mammalian olfactory G-protein-coupled receptors that biology uses, the readout can be expected in principle at least to eventually become biologically auditable: already we are starting to see the resolution necessary to be able to reconstruct which receptor was activated, when, and by how much. At present resolution we are catching only a faint whiff, but it is unmistakable: the scent of cancer can be captured by AI-enabled machines encoding the responses of stabilized GPCR olfactory receptors.

## Data Availability

he analysis is specified by the pseudocode and workflow flowcharts of SI-Code: a
pooled-control reference and feature fit on the training-partition controls only, an
L2-regularised logistic classifier without feature search, patient-grouped cross-validation,
whole-procedure label permutation, and patient bootstrap confidence intervals, with drift and
scaling assessed by leave-one-day-out, leave-one-chip-out, and learning-curve sub-sampling. All
reported values were computed on the single verified sensor export and its associated patient and
metadata records. Reasonable requests for the analysis code, result summaries, or de-identified
data will be considered, with priority given to academic collaborators, under appropriate
data-use and material-transfer agreements (contact)

## Supplementary Information

### SI-A. Analyses that did not survive their controls

*Each is an independently reproduced apparent result and the control that rejected it.* (a) Evolutionary in-sample handle selection: apparent ∼0.82, nested/permuted 0.49. (b) SCP random-handle search: best single-chip 0.83, whole-procedure permutation null 0.835 (*p* = 0.63). (c) a held-out batch’s collection-day proxy: pooled 0.79 collapses to 0.57 within-day (day predicted the label). (d) Within-cohort 0.79 versus cross-day 0.57 (cross-session drift; §8). (e) Mean-free-path density classifier: an anti-predictive AUC proved to be 1*/*density estimator instability (shuffled labels equally extreme). (f) Domain-adversarial “lift” of +0.06 on one seed vanished over five (*p* = 0.92). The general lesson: trust only the AUC number that survives permutation, bootstrap, and leave-one-*batch*-out.

### SI-B. The swarm / Machine Performance Index (biomimetic origin)

The starting estimator was a population of weak logistic learners, half reciprocally conditioned with cancer as the conditioned stimulus and half with control, averaged: a machine-learning version of the *Drosophila* olfactory Performance Index previously used with *Drosophila* by an author of this study [18–20], with its yoked control (labels shuffled) as the significance instrument. It is permutation-valid (LOPO 0.73, yoked-validated) but matched or beaten by plain L2-logistic, because reciprocal averaging cancels a common-mode offset that does not affect ranking (AUC). *The inspiration is the behavioural-assay architecture, reciprocal conditioning and the yoked null, not the vibrational/isotopic theory of olfaction; the method makes no claim about molecular-recognition mechanism*.

### SI-C. Estimators and signal codings evaluated

Estimators/recodings evaluated, all under the same protocol (permutation + patient-grouped CV): L2/L1-logistic, LDA, PLS-DA, SVM (linear/RBF), random forest, extra-trees, gradient boosting, *k*-NN, naive Bayes; reciprocal swarm/MPI; FlyHash; primacy-rank; curve-crossing shape; humidity residual/gain; concentration-ratio; MFP density; CORAL/quantile drift-removal; stacking; SPRT sequential sniffing; nested feature selection; noise augmentation. None exceeded L2-logistic on raw pooled-referenced magnitude features (0.79). Among the alternative signal codings, FlyHash sparse expansion reached 0.66 (a real but sub-logistic signal), primacy/onset-rank coding was at chance, and curve-crossing shape features were noise-dominated; each discards the multivariate relative-magnitude pattern and therefore underperforms.

### SI-D. Reproducibility

The analysis is specified by the pseudocode and workflow flowcharts of SI-Code: a pooled-control reference and feature fit on the training-partition controls only, an L2-regularised logistic classifier without feature search, patient-grouped cross-validation, whole-procedure label permutation, and patient bootstrap confidence intervals, with drift and scaling assessed by leave-one-day-out, leave-one-chip-out, and learning-curve sub-sampling. All reported values were computed on the single verified sensor export and its associated patient and metadata records. Reasonable requests for the analysis code, result summaries, or de-identified data will be considered, with priority given to academic collaborators, under appropriate data-use and material-transfer agreements (contact).

### SI-E. Reporting standard (STARD)

This study is reported in accordance with the STARD 2015 guidance [23] for diagnostic-accuracy studies, adapted for a retrospective, single-batch feasibility benchmark. The index test is the RealNose GPCR-MZI urine-headspace scent readout; the target condition is biopsy-confirmed prostate cancer; the reference standard is histopathological (biopsy) diagnosis. Participants were a convenience sample of biobanked specimens from men undergoing urological investigation at a single NHS hospital; no prospective enrolment, blinding of index-test operators to reference status beyond de-identification, or pre-specified clinical cut-point was applied. Discrimination is reported as area under the ROC curve with patient-level bootstrap confidence intervals and label-permutation significance; sensitivity/specificity at fixed operating points are reported where relevant. Indeterminate results are handled by the conformal-abstention analysis.

**Table SI-1:** STARD 2015 checklist, mapped to this report. “n/a” items reflect the retrospective single-batch feasibility design and are itemised as limitations.

| STARD 2015 item | Where addressed / status |
| --- | --- |
| Title/abstract: study as diagnostic accuracy | Title subtitle; structured abstract |
| Study objectives / hypotheses | Abstract (Objectives); §1 |
| Study design (prospective/retrospective) | Retrospective, single-batch; “Study at a glance” box, §2 |
| Setting and dates | Single UK NHS urology service; specimens biobanked $-80^{\circ}\text{C}$, 6 to 10 y (§2) |
| Eligibility / sampling of participants | Convenience biobank sample; Fig. SI-2, Table 1 |
| Index test (rationale, execution) | GPCR-MZI scent readout; §2 (detector), §3 (features/estimator) |
| Reference standard (rationale) | Biopsy/clinical diagnosis; §2 |
| Blinding of readers | Operators de-identified to reference status (no separate blinding); §2, this SI |
| Rationale for sample size | Feasibility benchmark (not powered); prospective power calc §?? |
| Statistical methods (estimates, precision) | Patient-grouped CV, AUC, bootstrap CI, permutation $p$; §3 |
| Handling of indeterminate results | Conformal-abstention analysis; §4 |
| Participant flow (STARD diagram) | Fig. SI-2 |
| Baseline clinical/demographic characteristics | Table 1 |
| Cross-tabulation / accuracy estimates | AUC and operating points; §4, Table 2 |
| Adverse events from index test | None (ex-vivo headspace only); “Biosafety” |
| Concordance / test variability | Test-retest disagreement 40%; repeatability §8.1 |
| Limitations | Discussion (Strengths and weaknesses), §12 |
| Registration / protocol | Retrospective; prospective protocol pre-specified §?? |
| Data availability | “Data and code availability” statement; version-controlled export |

### SI-E2. QUADAS-2 risk-of-bias and applicability summary

The STARD guideline defines which elements of diagnostic accuracy studies should be reported, whereas QUADAS-2 provides a framework for assessing risk of bias and concerns regarding applicability. Accordingly, we provide an explicit QUADAS-2 self-assessment (Table SI-2).

In line with the retrospective, single-batch feasibility design, we judged the patient selection domain to be at *high* risk of bias and to present *high* concerns regarding applicability. This rating is driven by the use of a non-consecutive biobank sample, a case–control design, and the absence of age-matching. Age-matching is an explicit design feature of the prospective study in the planning stages now but was not implemented in the present feasibility investigation.

We rated the index test domain as having *unclear* risk of bias. Test operators were de-identified with respect to reference-standard status rather than being formally blinded, and the operating threshold was determined by cross-validation instead of being prespecified *a-priori*.

We report these judgments explicitly rather than leaving them to be inferred, as they are intrinsic features of the feasibility-study design, are disclosed elsewhere in the manuscript, and are precisely the limitations that a prospective protocol currently bein designed is intended to address.

**Table SI-2:** QUADAS-2 summary. RoB = risk of bias. Ratings reflect a feasibility-study self-assessment; the accompanying basis cites where each is already disclosed.

| Domain | RoB | Applicability | Basis / existing disclosure |
| --- | --- | --- | --- |
| Patient selection | High | High | Non-consecutive convenience biobank sample with case/control selection; symptomatic work-up controls (not a screening population); any healthy-volunteer controls in the upstream programme self-identified and were not approached as patients were (Fig. SI-1), a differential-recruitment source of selection bias; cohort *not* age-matched (cases 67.7 y vs controls 51.8 y). Disclosed: “Study at a glance”, §2, Table 1, Fig. SI-1, Discussion, §?? (age-matched prospective design). |
| Index test | Unclear | Low | Index-test operators de-identified to reference status rather than formally blinded; decision threshold cross-validated, not pre-specified. Disclosed: Methods, Table 2, SI-E. |
| Reference standard | Unclear | Low | Cancers biopsy/histopathology-confirmed (accepted standard); controls defined by clinical work-up, which could miss occult disease. Disclosed: §2, label-limits. |
| Flow and timing | High |  | Reference basis differs between arms; specimens biobanked 6 to 10 y between collection and index measurement; all 80 labelled patients (40/40) are analysed, the discriminant being confined to the early analyte phase (§3). Disclosed: §2, Fig. SI-2. |

Upstream recruitment (context for patient selection). The analysed sensor cohort is a subset of a larger institutional recruitment programme at MKUH, whose STARD recruitment flow is reproduced in Fig. SI-1: of 1110 patients assessed, 901 consented and 892 entered the programme analysis, alongside 260 consented healthy volunteers (257 analysed). Two features are relevant to QUADAS-2 patient selection. First, healthy volunteers *self-identified and volunteered*, so they were not approached in the same way as patient participants, i.e. the two streams were recruited differentially. Second, the present study does *not* use this full programme cohort: to hold the measurement instrument fixed we analyse only the verified sensor subset (80 patients, 368 runs; Fig. SI-2), of which the controls are predominantly the symptomatic urological work-up population rather than these healthy volunteers. The recruitment flow is provided for transparency and does not correspond one-to-one to the analysed cohort.

**Figure SI-1:**
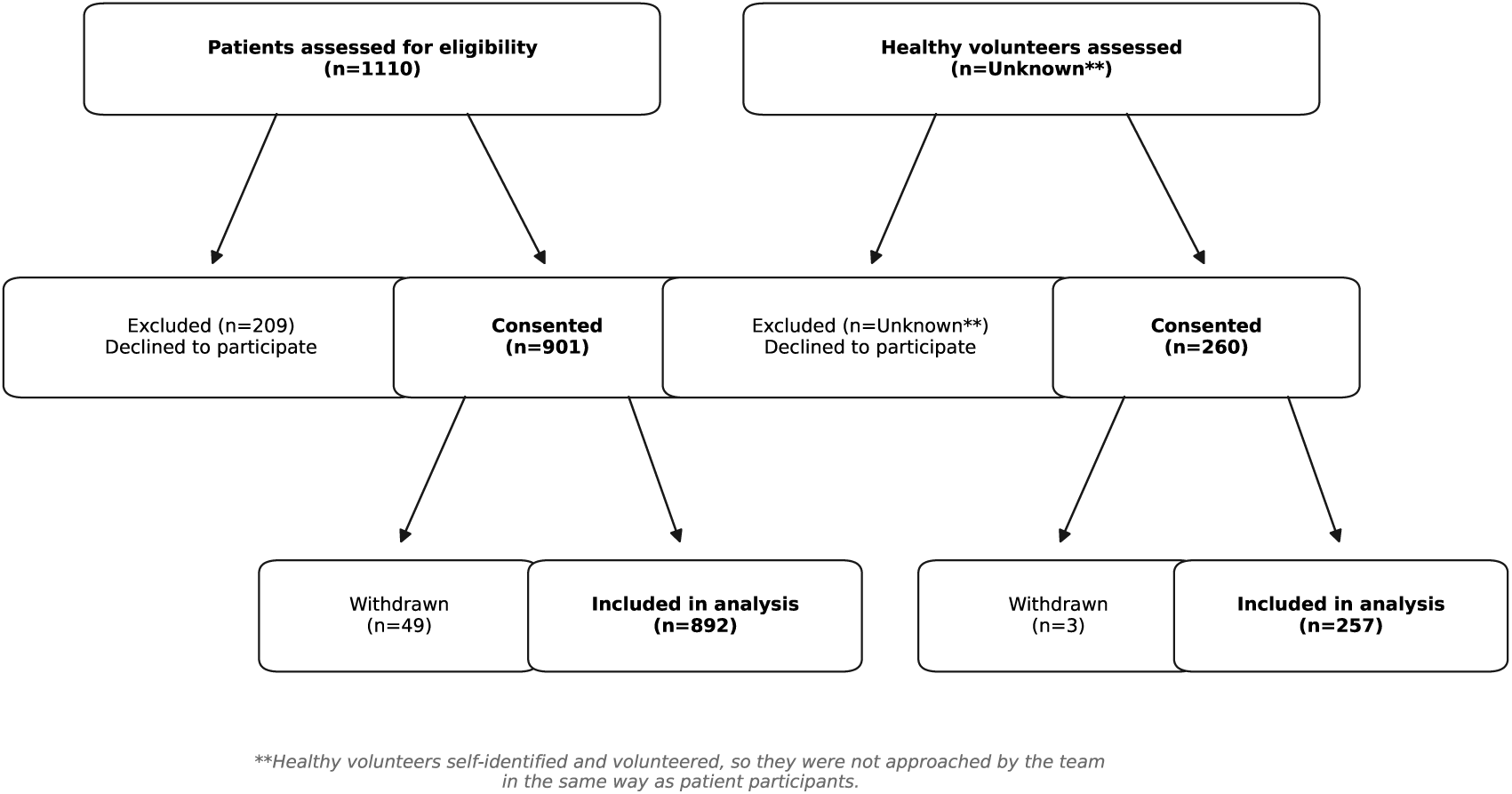
Upstream MKUH prostate-programme recruitment (STARD). Institutional recruitment of patients (1110 assessed → 901 consented → 892 analysed) and self-referred healthy volunteers (260 consented → 257 analysed). Healthy volunteers self-identified and were not approached as patients were. This documents the source programme; the present manuscript analyses the verified sensor subset of Fig. SI-2, not this full cohort.

**Figure SI-2:**
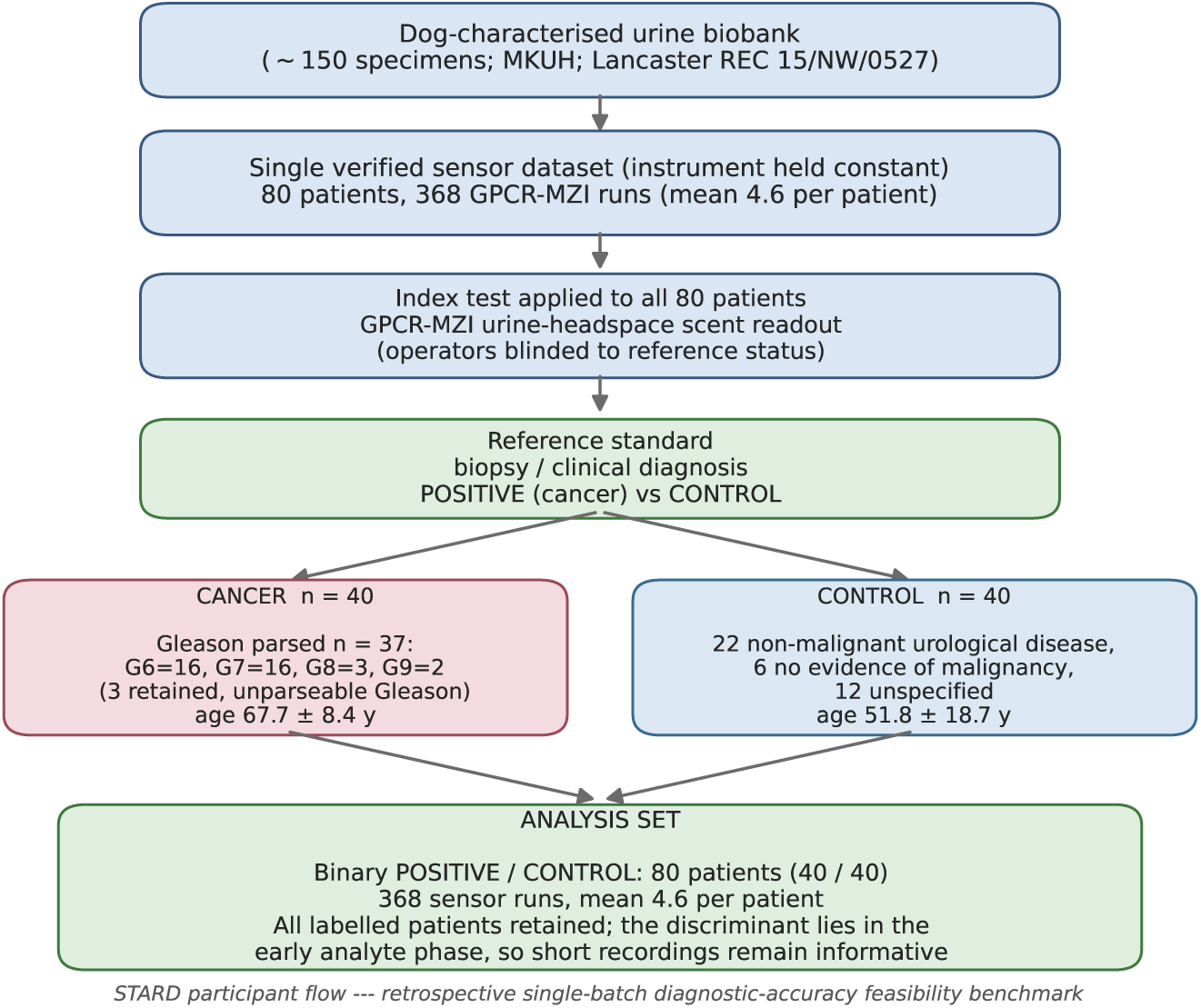
STARD participant-flow diagram for the analysed cohort. From the hosptial and dog-characterised urine biobank to a single verified sensor dataset (80 patients, 368 runs) held constant to isolate cross-instrument drift, through the index test (GPCR-MZI scent) and reference standard (biopsy/clinical diagnosis), to the analysis set: the binary POSITIVE/CONTROL cohort of 80 patients (40/40, 368 runs). All labelled patients are retained; because the discriminant lies in the early analyte phase, specimens whose acquisition stops before the late phase remain informative and are not excluded. Additional verified sensor datasets and legacy batches are held out to fix the instrument.

### SI-F. Neither age nor disease burden leaves a detectable scent trace, but prostate-cancer status does

We asked whether patient-level attributes other than the binary label leave a readable trace in the scent readout, using the fold-honest leave-one-patient-out (LOPO) score merged with the clinical workbook (Gleason sum, positive-core fraction, PSA, risk, final diagnosis; study cohort). Three findings, all single-batch and hypothesis-generating:

#### (i) Disease burden is not encoded

The model score is uncorrelated with any burden variable, Gleason sum *ρ* = −0.07 (*p* = 0.68, *n* = 37), positive-core fraction *ρ* = 0.05 (*p* = 0.78), and PSA *ρ* =−0.15 (*p* = 0.38, *n* = 38). Higher-grade or higher-volume disease is not easier to detect.

#### (ii) Comorbid disease does not raise a generic cancer scent

No control in this cohort carries a documented cancer (all 40 are non-malignant disease, no-evidence, or unspecified), so the literal “other cancers elevate cancer scent” hypothesis is untestable here. Using non-malignant *disease* vs *no evidence of disease* as a graded proxy, disease controls were scored *lower* (mean 0.39, false-positive rate 0.27) than disease-free controls (mean 0.50, FP 0.50; Mann-Whitney *p* = 0.24, *n* = 22 vs 6), the opposite of the hypothesised direction, and underpowered.

#### (iii) Total signal magnitude is near-chance and burden-independent

Overall response amplitude discriminates cancer at only AUC 0.54 (matching the total-magnitude scalar of §5), does not track burden (Gleason *ρ* = 0.03, cores *ρ* = 0.01, PSA *ρ* =−0.21), and only weakly co-varies with the model score (*ρ* = 0.22, *p* = 0.045). A faint control-only thread (PSA vs magnitude *ρ* = 0.47, *p* = 0.06, *n* = 16) is noted for larger cohorts.

Taken together, the discriminant is specific to prostate-cancer *status* as a present/absent olfactory character; it does not read age (§6), disease burden, comorbidity, or intensity. This is reassuring for specificity and consistent with the paper’s framing that the dominant factor is training-set size, not disease severity.

#### Receptor-set ablation

On the eight receptors present on the chip, LOPO on the scale-free handle descriptors gives: all eight = 0.67; the potentially prostate-relevant subset (hOR51E1, hOR51E2/PSGR, hOR17-4, hOR2H1) = 0.59; the remaining four (hOR2AG2, hOR2T7, hOR5M10, hOR8G5) = 0.61; and no single receptor exceeds 0.53 alone (PSGR pair 0.52). No receptor subset dominates: discrimination is a distributed pattern that improves with receptor breadth (see also §5).

### SI-G. Chip protein population, spot placement, and why aggregation is by receptor identity (not by spot)

The measurement hardware is not uniform across chips, and disambiguating this is essential to how features are formed. Two chip builds appear in the analysed sensor export (Table SI-3). Both share the same 64-spot silicon-photonic Mach-Zehnder interferometer geometry [5]; only the receptors printed onto those 64 spots differ. Batch 1 carries *four* receptors, each printed at *two* concentrations a factor of ∼6 apart (2 of 6 chips: UYEC7, YAG99). Batch 2 carries *eight* receptors, each at a *single* concentration: the same four as batch 1 plus four additional receptors (4 of 6 chips: 4GK8A, DNS33, F8TE4, U9572). The full protein population and its concentrations are listed in Table SI-4. The only receptors common to *every* chip (and to the other cohorts, e.g. the four-receptor single-concentration LXKNX build in another cohort) are the four batch 1 receptors at their high concentration: hOR17-4@4.73, hOR51E1@10.22, hOR51E2@12.48, hOR2H1@16.70; cross-chip and cross-cohort analyses therefore lean on these four.

Same spot, different receptor. Crucially, the physical grid layout is reshuffled between builds: the identical grid cell hosts a *different* receptor depending on the build (Table SI-5). Averaging signals by *spot placement* across chips would therefore pool chemically distinct receptors (e.g. hOR51E1 with hOR17-4). We consequently key every feature by receptor identity, the (protein, concentration) pair, and pool over a receptor’s physical replicate spots *and* over the batch 1 and batch 2 code schemes that denote the same (protein, concentration).

Concentration normalisation. On the batch 1 dual-concentration receptors, the ∼6-fold concentration step yields a ∼2-fold signal step (ratios 5.94 to 6.01×; *k* = log_6_ 2 = 0.39; §5), which we use both as a receptor-mediated-transduction control and as a built-in per-spot quality check. This dual-concentration design exists only on the batch 1 build; the batch 2 receptors are single-concentration and do not carry the internal ratio QC.

**Table SI-3:**
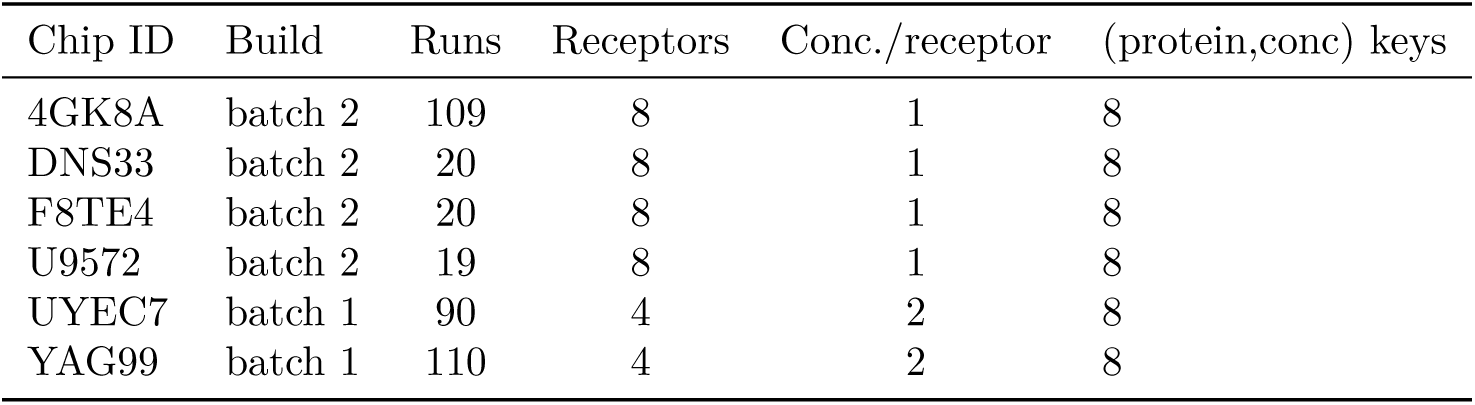
Chip protein population in the analysed sensor dataset (368 runs, 6 chips). “keys” = distinct (protein, concentration) feature keys.

**Table SI-4:**
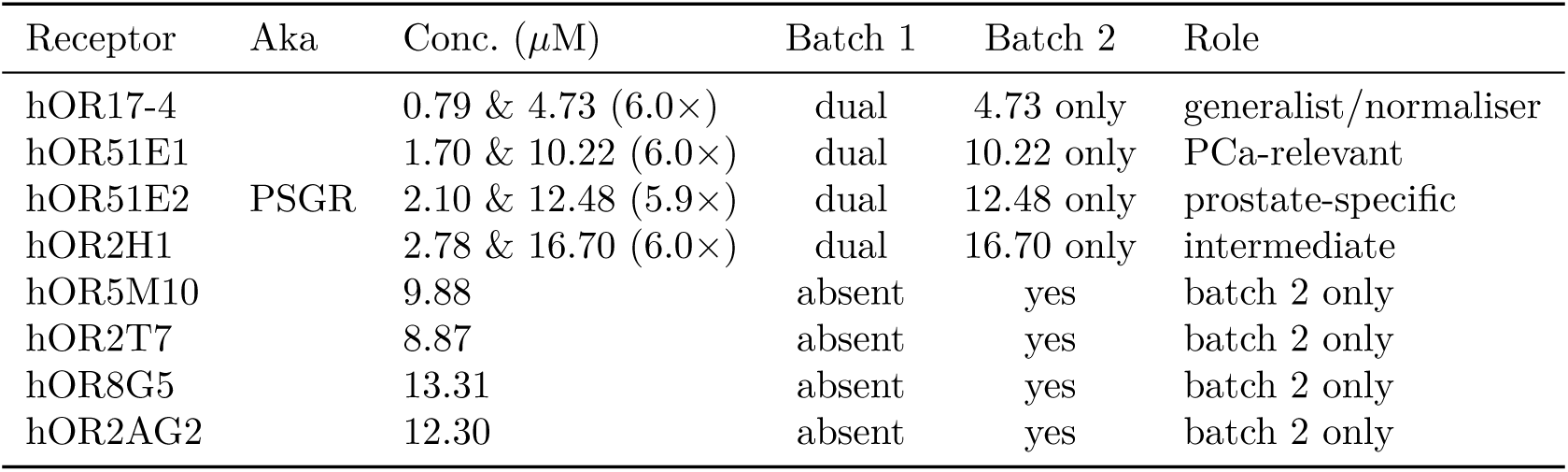
Full receptor population and concentrations. Batch 1 prints each of the four at two concentrations (∼6× apart); batch 2 prints eight at one concentration each. Codes 1 and 10 are reference/blank spots (excluded).

**Table SI-5:** Spot-placement collisions: the same physical grid cell (four replicate columns per row letter) carries a *different* receptor in the two builds, hence aggregation must be by receptor identity, not by spot.

| Grid cell (rows) | Batch 2 receptor | Batch 1 receptor |
| --- | --- | --- |
| B0 to B3 | hOR51E1@10.22 | hOR17-4@4.73 |
| C0 to C3 | hOR51E2@12.48 | hOR51E1@10.22 |
| D0 to D3 | hOR2H1@16.70 | hOR51E2@12.48 |
| E0 to E3 | hOR5M10@9.88 | hOR2H1@16.70 |
| F0 to F3 | hOR2T7@8.87 | hOR17-4@0.79 |
| G0 to G3 | hOR8G5@13.31 | hOR51E1@1.70 |

### SI-H. Detector mechanism: receptor-mediated transduction and dose-response

A recurring worry for any optical biosensor is that the measured signal reflects a bulk or instrument effect, refractive-index shifts, non-specific adsorption, humidity, or optical drift, rather than receptor-ligand binding. Three independent lines of evidence show the response is due to the receptor protein itself. *(i) Receptor-dose dependence.* Each of the four dual-concentration receptors is printed at two concentrations a factor of ∼6 apart; the analyte response scales up by 2.00 ± 0.14 (range 1.86 to 2.19; CV 7%) with the ∼6-fold increase in printed receptor. A bulk or instrument artifact is independent of how much receptor occupies a spot and would give a ratio near unity; the strong, monotone dependence on receptor loading therefore places the response in the receptor. *(ii) Saturable, not linear.* The scaling is compressive, with a common exponent *k* = 0.39 ± 0.04 (activation ∝ *C^k^*; log_6_ 2 = 0.387), the Langmuir/Hill signature of *saturable specific binding*, whereas non-specific bulk effects scale approximately linearly (*k* ≈ 1). *(iii) Receptor-general constancy.* This ∼2×-per-6× law holds to within a few percent across all four chemically distinct receptors (*p* ≈ 10*^−^*^34^; *n* = 368), indicating a single systematic receptor-mediated transduction mechanism rather than idiosyncratic per-spot contamination. Together with the MZI dual-path cancellation of bulk humidity/temperature (both interferometer arms sample the same ambient) and the independence of the diagnostic signal from humidity, temperature, collection day, age and overall amplitude (§6), this establishes the discriminant as receptor-borne olfactory transduction. As a further control the concentration *ratio* carries no cancer information (AUC 0.50), localising the diagnostic signal to the cross-receptor pattern rather than the dose-response magnitude. The dual-concentration design additionally furnishes a built-in per-spot quality check: because the high/low ratio is so tightly conserved (inter-quartile spread ±2 to 10%), a spot pair falling outside a narrow band flags a denatured, defective or fouled receptor without any external calibrant. This dual-concentration design exists only on the batch 1 build (SI-G).

**Figure SI-3:**
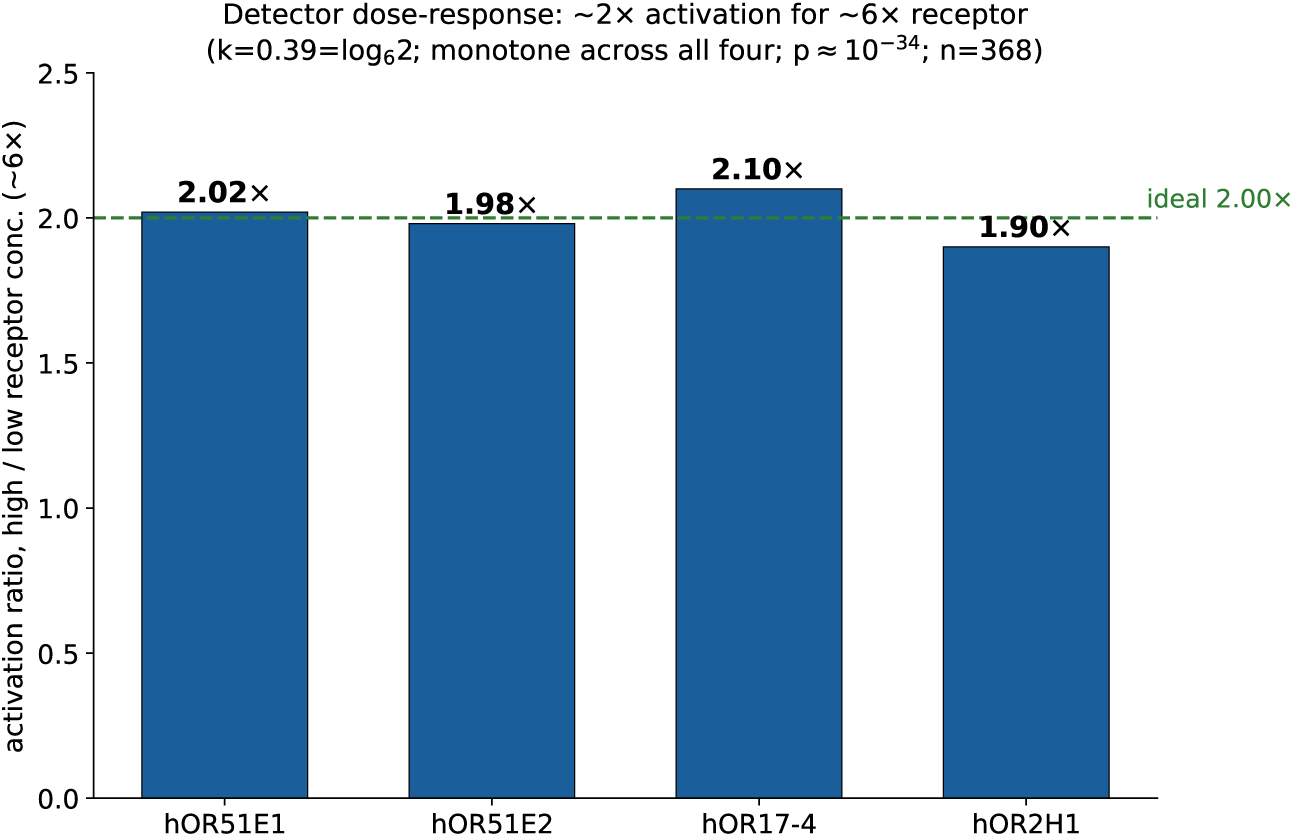
Detector dose-response check: same-protein high-versus low-concentration spots activate ∼1.9 to 2.2× (sub-linear; monotone across all four receptors; *p* ≈ 10*^−^*^34^; *n* = 368). The detector is a saturating binder, but the saturation ratio itself carries no cancer information (AUC 0.50; §5), localising the discriminant to the cross-receptor pattern.

### SI-Code. Pseudocode and workflow flowcharts

The complete analysis pipeline is specified below in human- and machine-readable pseudocode, sufficient for an independent group to reproduce the reported values from the verified sensor export, with the corresponding workflow flowcharts in Figs. SI-4 to SI-7.

#### Feature construction and fold-honest referencing

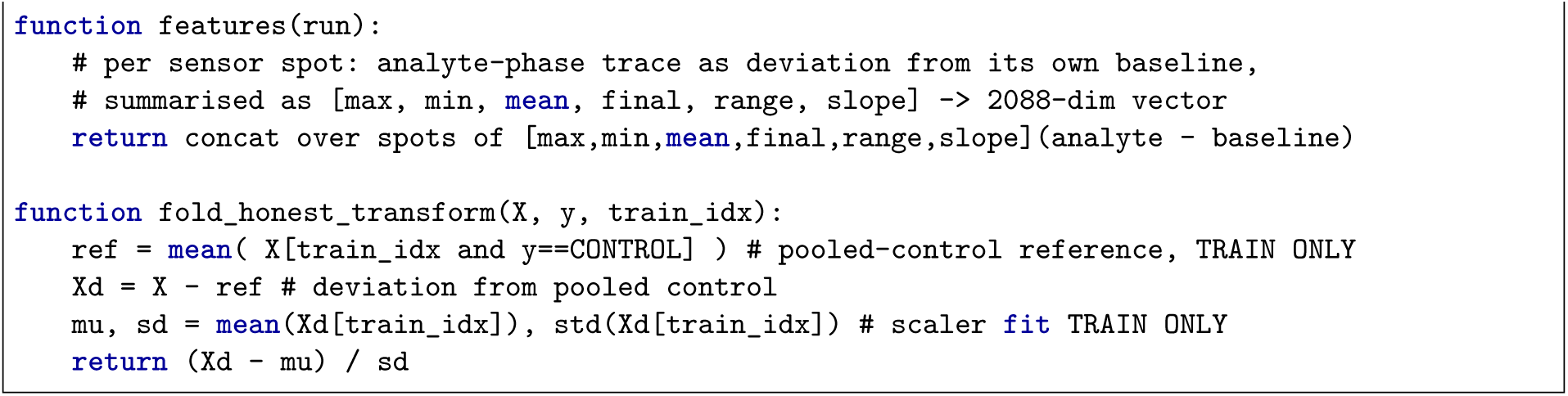

#### Primary estimator and patient-level evaluation

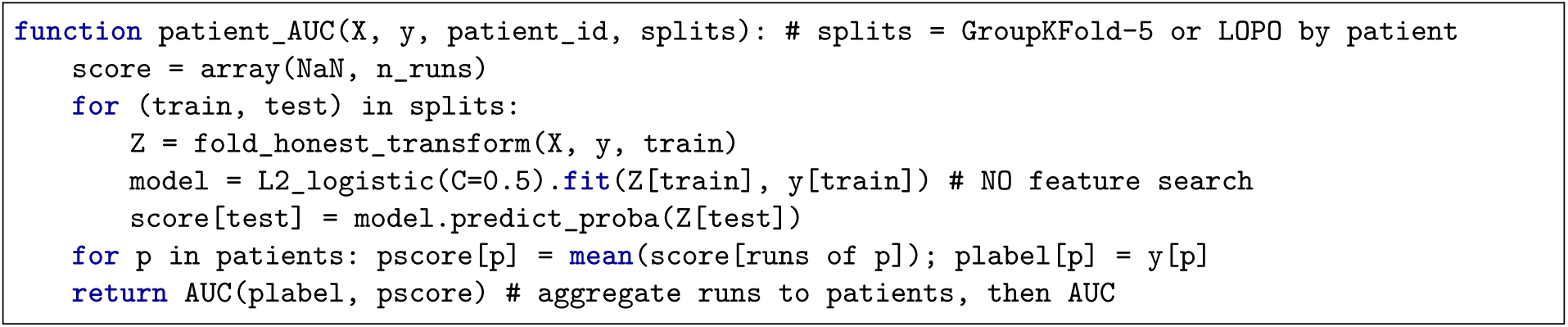

#### Significance and uncertainty

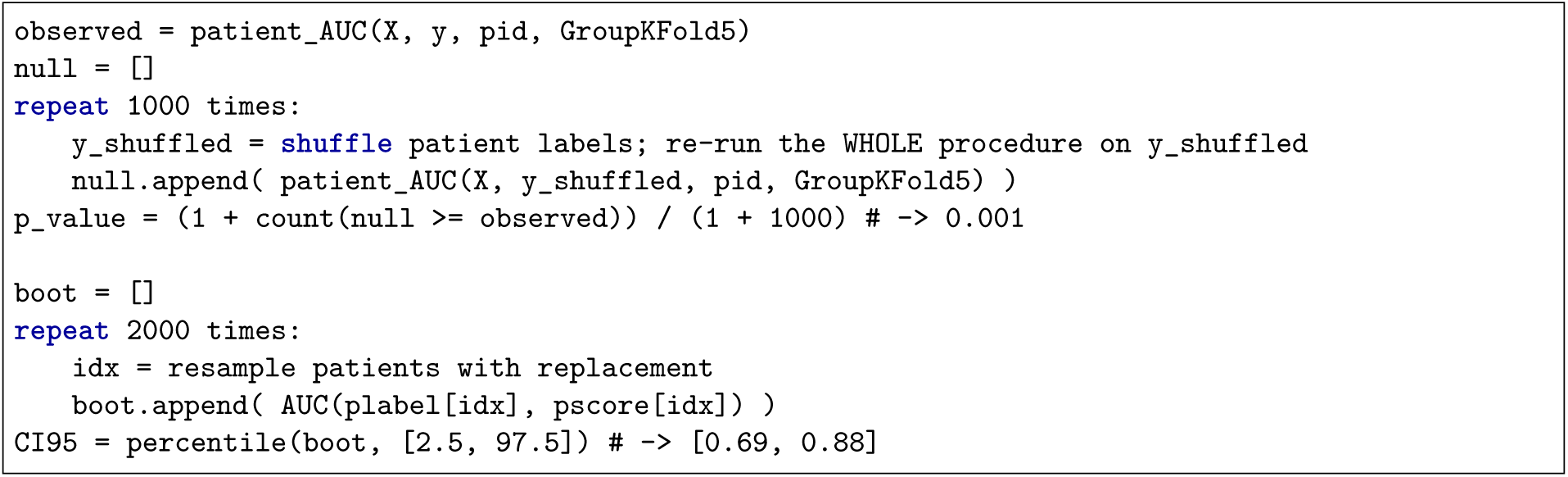

**Figure SI-4:**
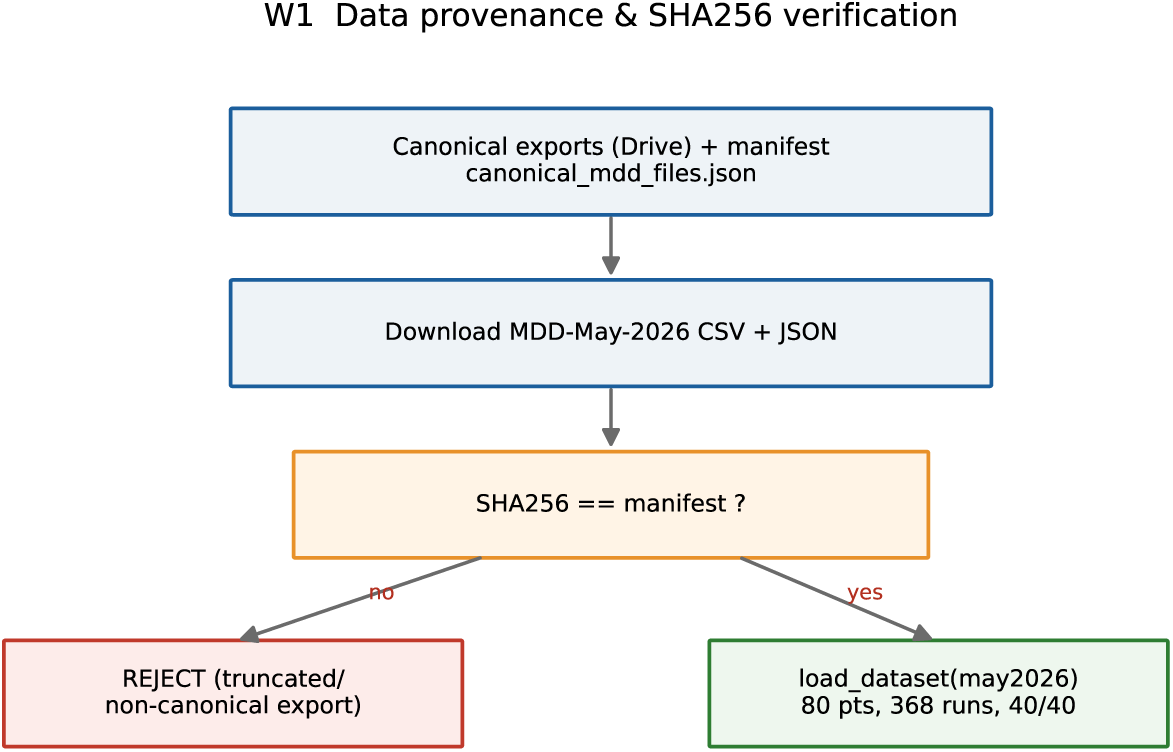
Workflow W1: data provenance and verification.

**Figure SI-5:**
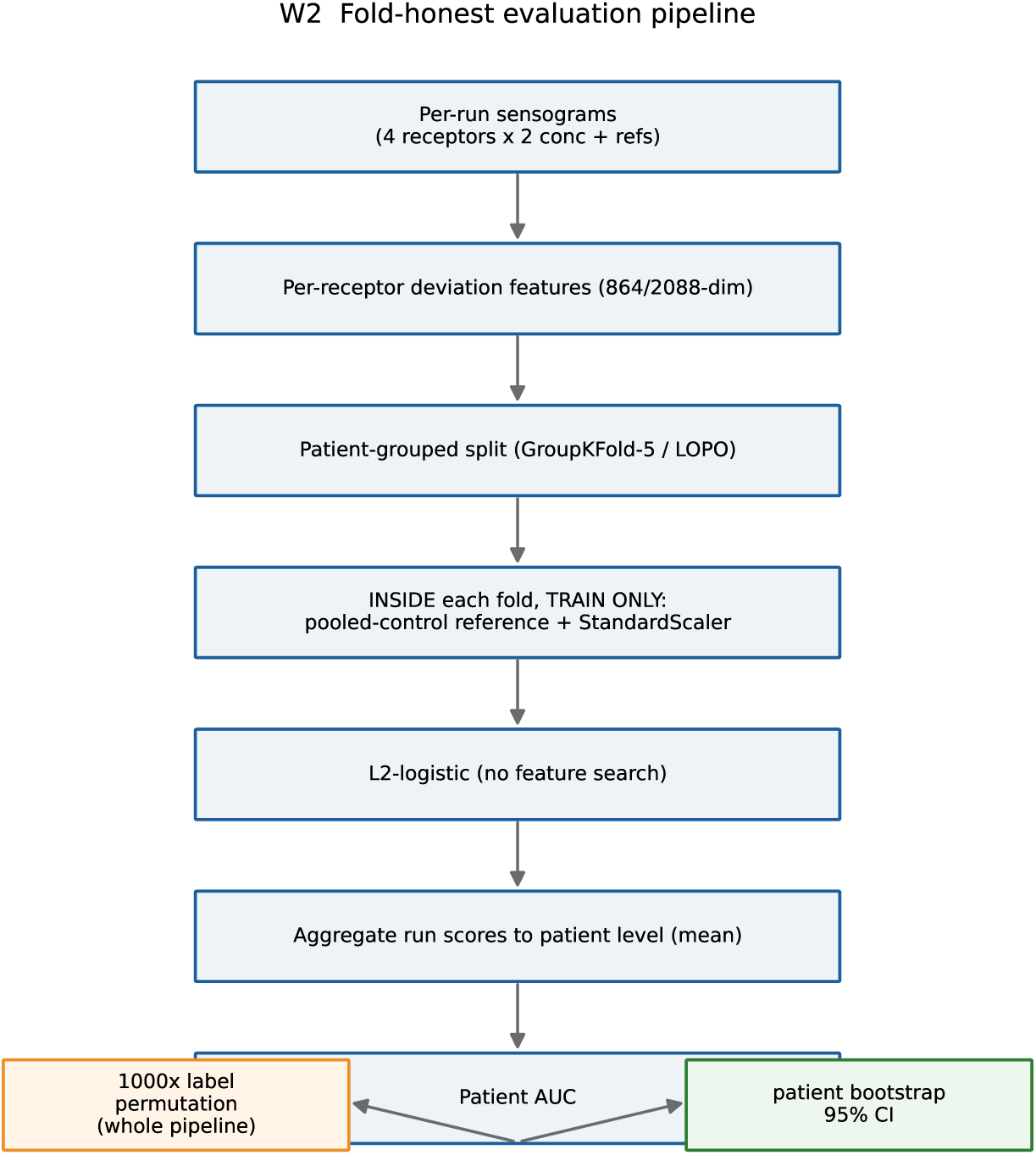
Workflow W2: fold-honest evaluation and significance.

**Figure SI-6:**
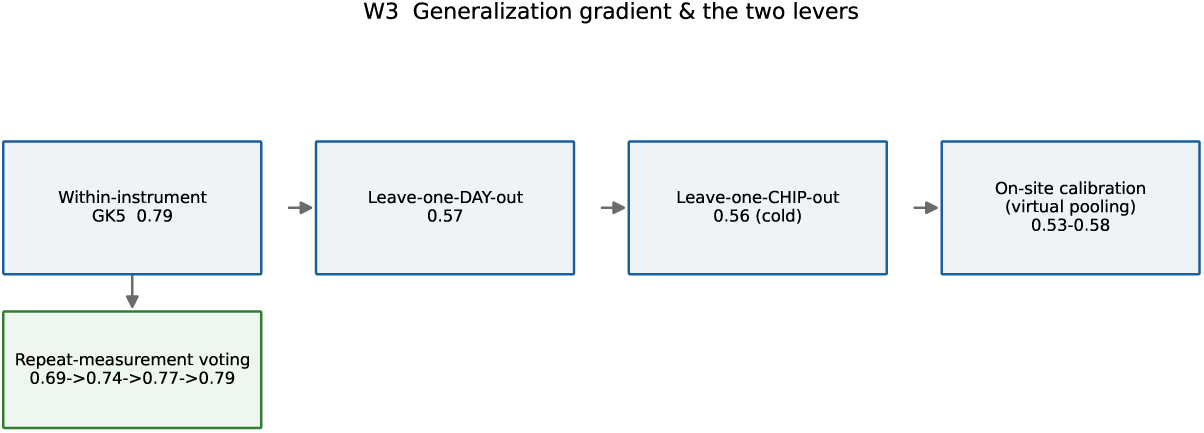
Workflow W3: generalisation gating and on-site calibration.

**Figure SI-7:**
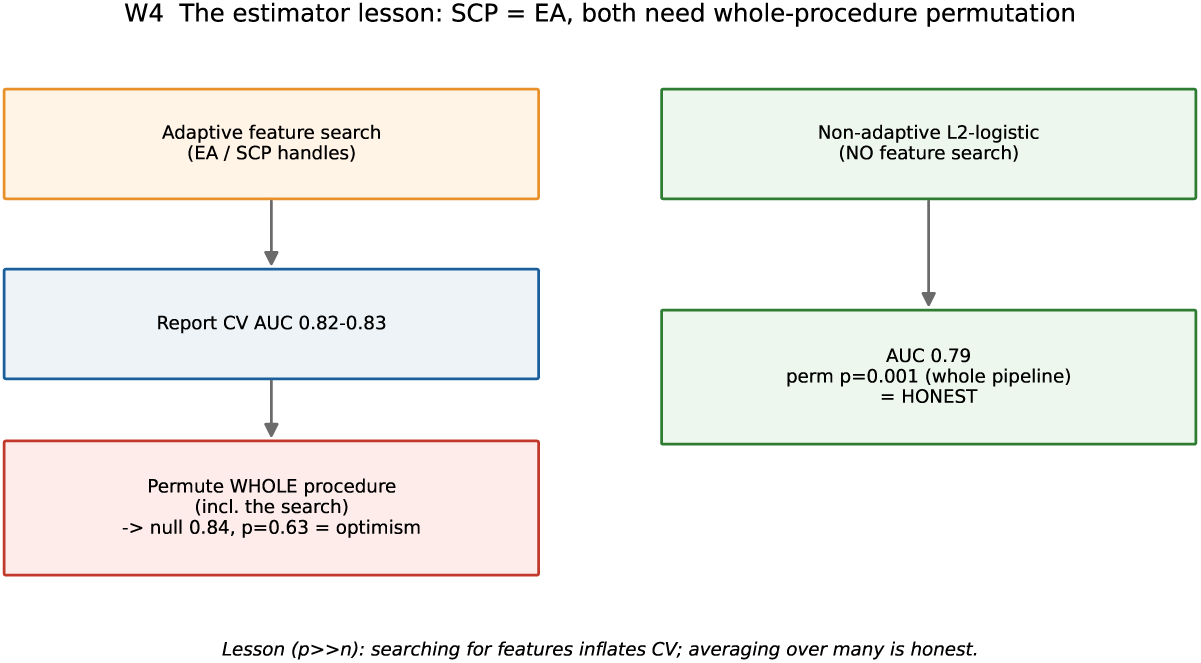
Workflow W4: the estimator lesson (adaptive search versus averaging).

## Ethics, human subjects and privacy

The canine work that inspired this study followed the ethical and welfare standards specified by Medical Detection Dogs, with handler and institutional oversight. The present study was approved by the Lancaster Research Ethics Committee, North West, United Kingdom (Ref: 15/NW/0527). All participants provided written, informed consent that explicitly covered the de-identified use of biospecimens and associated metadata for secondary research, method development, and controlled sharing with external investigators under data-use/material-transfer agreements (DUAs/MTAs) and local approvals. To protect privacy, shared datasets were stripped of direct identifiers (name, NHS number, full postal address, exact date of birth); where dates are required for analysis they are reported in relative form (days from collection) or aggregated (month/year). Access to de-identified data and analysis code is considered by a Data Access Committee under a standard DUA prohibiting re-identification and redistribution. Sharing complies with applicable data-protection law (UK GDPR; HIPAA as applicable) and institutional policy.

## Biosafety and regulatory transparency

The RealNose platform uses purified, stabilised mammalian olfactory G-protein-coupled receptors expressed in HEK293 cells for upstream production. *No live mammalian cells or replicative biological agents are present in the finished sensor chips used for measurement*: purification and immobilisation include inactivation and clearance steps and yield a non-viable, proteinaceous surface. Cell culture and receptor expression were performed under appropriate containment (BSL-2 where required) and Good Laboratory Practice or equivalent; chip functionalisation and assembly were performed in controlled clean-room environments with documented quality control for lot consistency. For the study reported here the device samples only the ex-vivo headspace above an aliquot of urine; there is no patient contact with immobilised GPCRs or sensor internals. Consumables and reusable components that contact sample headspace follow validated cleaning, sterilisation and disposal workflows. Manufacture and commercialisation are planned under Good Manufacturing Practice; planned regulatory and safety activities include analytical verification (accuracy, precision, reproducibility across lots and sites), formal stability and shelf-life testing, ISO 10993 biocompatibility testing for any future patient-contacting elements, and device risk management under ISO 14971.

## Competing interests

AM is a co-founder, employee and equity holder of RealNose, Inc. (RNI) and a named inventor on patent applications and provisional filings relating to machine olfaction and GPCR engineering; CG (Guest) is a co-founder and employee of Medical Detection Dogs (MDD), which provided programmatic support and the urine samples used here, and declares no personal equity stake in RNI; NS is a co-founder, employee, equity holder and named inventor; RH and CD are employees of MDD with no personal equity in RNI; AR is a contractor to RNI, founder of Hcyon Technology and a named inventor on debiasing/machine-olfaction filings; SJ and AK (Kotsis) are consultant/contributor to RNI; KCK is an unpaid intern at RealNose; ZK is an employee of RNI; HK is affiliated with Endless Frontiers Laboratory / NYU Stern and provides advisory support with equity interest; EZ is employed by Cleveland Clinic and provides advisory support with equity interest; CG (Gluck) directs Dr. Gluck’s Wellness Center and serves as an equity-holding advisor to RNI; IA, FT, AC and TL are employed by Milton Keynes University Hospital NHS Foundation Trust and contributed clinical / sample resources with no personal financial interest in RNI; SZ, KO and PPL are affiliated with MIT and received institutional support with no personal equity interest in RNI. RNI provided financial and in-kind support including sensor hardware, reagents and engineering resources. The authors confirm these interests do not alter adherence to standards on data and material sharing.

## Acknowledgments

We thank RealNose bioengineering staff and interns for data collection; the staff, volunteers and board of Medical Detection Dogs (UK) for canine detection and oncology insight; Dr. Alan Makepeace for helpful discussions; the Gluck Wellness Center for early lab-workflow support; the olfaktion.com/Neose creators (with particular thanks to Dr. Tristan Rousselle and Dr. Cyril Herrier) for the MZI chip; FTL-INC (Nancy Kasberg) for figure production; Cube Biotech (Dr. Maryna Lowe, Dr. Barbara Martens, Dr. Jan Kubicek) for receptor expression and stabilisation; and colleagues at MIT Lincoln Laboratory and the MIT Media Lab. We gratefully acknowledge financial support and advice from Glenn Krevlin (GlenHill Capital), Nelson Wang and George Kung (KungHo Fund), and Armando Cañas and Robert Jursich (Mavericus Fund), the NATO Defence Innovation Accelerator for the North Atlantic (DIANA) programme, and support from the U.S. Department of War, and prior support from DARPA, GlaxoSmithKline and the Prostate Cancer Foundation.

## Data and code availability

Reasonable requests for the de-identified measurement data, model code, and detailed protocols will be considered, with priority given to academic collaborators, under appropriate data-use and material-transfer agreements and institutional-review-board permissions (contact). All results in this paper were computed on a single, version-controlled sensor export; because a column-truncated export with the same 80 patients inflates the reported value (0.79 → 0.82), the exact file version must be used, and a checksum is provided with the data on request.

## Glossary

*Written for a clinical reader; each machine-learning or statistical term is defined in plain language, with a clinical analogy where one helps*.

### Clinical, sample and study terms

**BPH** Benign prostatic hyperplasia, non-cancerous prostate enlargement that itself raises serum PSA.

**Control** In this study, a patient whose reference test showed no prostate cancer (mostly other, non-malignant urological disease); *not* a healthy volunteer.

**DRE** Digital rectal examination.

**Gleason score / sum** The histological grade of prostate cancer from biopsy (two patterns summed, e.g. 3+4=7); higher = more aggressive.

**Positive-core fraction** Of the biopsy needle cores sampled, the proportion containing cancer, a rough measure of tumour volume/extent.

**Index test** The test being evaluated (here, the RealNose urine-scent readout).

**Reference standard** The “truth” a diagnostic test is judged against (here, biopsy / clinical diagnosis).

**Prevalence** The fraction of the tested group who actually have the disease (50% here by design; it changes PPV/NPV, not sensitivity/specificity).

**Sensitivity** Of patients *with* cancer, the fraction the test calls positive (true-positive rate).

**Specificity** Of patients *without* cancer, the fraction the test calls negative (true-negative rate).

**PPV / NPV** Positive / negative predictive value, given a positive (negative) test, the probability the patient truly does (does not) have cancer, at the stated prevalence.

**Youden-optimal point** The score threshold that maximises sensitivity + specificity −1; a common “balanced” operating point.

**TNM** Standard tumour-node-metastasis staging.

**REC / IRB** Research Ethics Committee / Institutional Review Board, the bodies that approve human-subjects research.

**STARD** Standards for Reporting of Diagnostic Accuracy Studies, the reporting checklist this paper follows.

**MDD / MKUH** Medical Detection Dogs / Milton Keynes University Hospital.

**PCa** Prostate cancer. PSA Prostate-specific antigen (the standard blood test). csPCa Clinically-significant prostate cancer. mpMRI Multiparametric MRI.

### Sensor, chemistry and signal terms

**GPCR** G-protein-coupled receptor, the large family of cell-surface proteins that includes the human smell receptors used here as the sensing element.

**OR / olfactory receptor** A GPCR that binds odour molecules; e.g. hOR51E1, hOR51E2.

**PSGR** Prostate-specific G-protein-coupled receptor (the receptor OR51E2), expressed in prostate tissue.

**HEK293** A standard laboratory human cell line used to manufacture the receptor proteins.

**MZI** Mach-Zehnder interferometer, an optical chip that senses tiny changes when an odour binds a receptor by comparing light along two paths (which also cancels shared temperature/humidity effects).

**VOC** Volatile organic compound, an airborne small molecule; the “smell” chemicals in urine headspace.

**Headspace** The air above a liquid sample that carries its volatile molecules to the sensor.

**Analyte phase / sensogram** The time-trace of the sensor’s response as the sample air (“analyte”) passes over it.

**Spot / grid cell** One printed micro-deposit of receptor on the chip; multiple replicate spots per receptor.

**Pooled-control reference / virtual pooling** The average sensor response of the non-cancer (control) samples, subtracted from every reading so each patient is expressed as a *deviation from normal urine*, the analysis analogue of running a batch of local control urines to calibrate a site.

**FFT (fast Fourier transform)** A standard maths operation that summarises the *shape* of a time-trace as a few numbers (its frequency components).

**Langmuir / Hill saturable binding** The classic chemistry of a receptor whose response grows less than proportionally as more odour (or more receptor) is added because binding sites saturate, the signature we use to prove the signal is genuine receptor binding, not a bulk artefact.

### Machine-learning and statistics terms

**Feature** One input number fed to the classifier (here, a summary of one receptor’s response); this study uses 2088 features per measurement.

**Classifier / model** A formula that combines the features into a single score (0 to 1) estimating the probability of cancer.

**Logistic regression** The simplest standard classifier: it takes a weighted sum of the features and converts it to a probability, essentially a multi-variable version of the risk scores clinicians already use.

**L2 regularisation (“ridge”)** A restraint added to logistic regression that keeps every feature’s weight small and shares influence across many features rather than letting a few dominate; it prevents the model from over-fitting noise and is why our model is a diffuse, “vote” across all receptors. L1 (“lasso”) is the alternative that instead forces most weights to exactly zero (picks a few features).

**Standardisation (StandardScaler)** Rescaling each feature to a common scale (mean 0, spread 1) so no single feature dominates merely because its units are larger.

**Feature selection** Choosing a subset of features before/while fitting; “adaptive” search over many subsets can look impressive in-sample but often does not generalise (see §6).

**Overfitting / optimism** When a model learns noise peculiar to the training patients and so reports a better score than it will achieve on new patients.

**Cross-validation (CV)** Repeatedly training on part of the data and testing on the held-out part, so every patient is scored by a model that never saw them.

**Patient-grouped CV / GroupKFold** Cross-validation that keeps *all* of a patient’s repeat measurements together on the same side of the split, so no run of a test patient leaks into training.

**LOPO / LODO / LOCO** Leave-one-patient-out / leave-one-day-out / leave-one-chip-out, progressively stricter versions that hold out one whole patient, one whole measurement day, or one whole sensor chip, to test generalisation to new patients, days, and hardware.

**Leakage / fold-honest** “Leakage” is any way test information sneaks into training and inflates the score; “fold-honest” means the reference and scaling are computed on the training patients only, so there is none.

**AUC (area under the ROC curve)** A single 0.5 to 1.0 summary of how well the score separates cancer from control across all thresholds: 0.5 is a coin-flip, 1.0 is perfect; it is the probability that a random cancer patient scores higher than a random control.

**ROC curve** The plot of sensitivity versus (1 − specificity) as the decision threshold is swept.

**Permutation test / *p*-value** Re-running the *entire* analysis many times with the cancer/-control labels randomly shuffled to see how often chance alone matches the real result; *p* = 0.001 means once in a thousand, i.e. the signal is very unlikely to be luck.

**Bootstrap / 95% confidence interval** Re-computing the AUC on many random resamples of the patients to get a plausible range; the 95% CI is the interval that contains the true value about 95% of the time.

**Learning curve** A plot of accuracy versus training-set size; its upward slope is our central point: like a dog or a trainee, the system improves with more examples.

**Inverse-power(-law) fit** The simple curve AUC(*n*) = *A* − *B n^−γ^* commonly used to describe how accuracy rises with sample size *n*, used here to extrapolate.

*p* ≫ *n* **regime** “More features than patients”, the statistically hard setting (here 2088 features, ∼80 patients) where over-searching is dangerous and averaging is safer.

**Conformal abstention / coverage** A rule that declines to call the least-confident cases (“coverage” = the fraction it does call), improving accuracy on the rest, akin to a clinician reporting an equivocal result as indeterminate.

**Spearman** *ρ* A correlation coefficient (−1 to +1) measuring whether two quantities rise and fall together, without assuming a straight-line relationship.

**Mann-Whitney *U* / point-biserial** Standard non-parametric / two-group statistical tests used here for small samples.

**Domain shift / drift; CORAL** “Drift” is the change in sensor behaviour between chips or days; CORAL is one standard method that tries to align the two, with only three days here it is under-determined.

**Cold vs calibrated transfer** “Cold” = a fixed model applied to a new site unchanged; “calibrated” = first re-referencing to a few local control urines (on-site acclimatisation).

**Swarm / Machine Performance Index (MPI)** A biologically-inspired ensemble of many small models, transcribing the reciprocal-conditioning “Performance Index” used in *Drosophila* smell assays; it survives its yoked null but does not beat plain logistic regression here (SI-B).

**SCP / EA** Self-calibrating-protocol handle search [6–8] / evolutionary algorithm, two adaptive feature-search methods that, being the same idea, inflate cross-validation identically and collapse under whole-procedure permutation (§6).

**FlyHash** An insect-inspired way of expanding features into a sparse code; tested and weaker than logistic regression here.

## Notes

### Author Declarations

Cohort and ethics. Urine was collected under Milton Keynes University Hospital governance (Lancaster REC 15/NW/0527) from men undergoing urological investigation, including biopsy- confirmed prostate cancer across a range of Gleason grades, benign prostatic hyperplasia (BPH), and age/symptom context controls; all gave informed consent. The present study was approved by the Lancaster Research Ethics Committee, North West, United Kingdom (Ref: 15/NW/0527).

